# Transmissibility of COVID-19 among Vaccinated Individuals: A Rapid Literature Review - Update #1

**DOI:** 10.1101/2021.06.29.21255526

**Authors:** Oluwaseun Egunsola, Liza Mastikhina, Laura Dowsett, Brenlea Farkas, Mark Hofmeister, Lynora Saxinger, Fiona Clement

**Affiliations:** The Department Community Health Sciences, Teaching Research and Wellness Building, 3280 Hospital Drive NW Calgary Alberta T2N 4N1; O’Brien Institute for Public Health, Teaching Research and Wellness Building, 3280 Hospital Drive NW Calgary Alberta T2N 4N1; Department of Medicine University of Alberta, Edmonton, Alberta, Canada

## Abstract

**Objectives:** This is an update of a previous report that examined literature published up to March 11th, 2021. Sixteen additional studies have been included in this update. The objective of this report is to identify comparative observational studies and randomized controlled trials (RCTs) evaluating the efficacy and effectiveness of COVID-19 vaccination in reducing forward transmission from vaccinated people, and studies examining the biological plausibility of vaccination-induced transmission reduction.

**Method:** A search of databases, MEDLINE, Embase, L-OVE and the Cochrane Central Register of Controlled Trials was conducted to identify RCTs or comparative observational studies evaluating the efficacy and effectiveness of COVID-19 vaccination in the prevention of transmission, asymptomatic infections and transmissibility of COVID-19 among vaccinated persons. An additional search of grey literature was conducted. This search is current to May 4th, 2021.

**Results:** In this update, 16 additional studies, including 9 human and 7 animal studies, were included. Therefore, this review examines a total of 33 included studies: 21 human studies and 12 preclinical animal studies. Evidence from two large household surveillance studies from the UK suggests that a single or full dose of AstraZeneca (AZ) and Pfizer-BioNtech (PfBnT) vaccines may prevent household transmission of COVID-19 after 14 days of vaccination by up to 54%. The AZ vaccine trials in the general population suggest that an initial low dose followed by a standard dose may provide up to 59% protection against asymptomatic or unknown infection, although efficacy against these outcomes was not demonstrated following two standard doses. PfBnT vaccine observational studies in the general population suggest up to 90% effectiveness against asymptomatic infection after seven or more days of full dose vaccination. Up to 75% effectiveness against asymptomatic infection was reported after full-dose in healthcare workers. Across RCTs examining asymptomatic infection in the general population, one dose of Moderna was shown to provide an efficacy of 61.4% against asymptomatic infection 21 days after the first dose; in another trial, the J&J vaccine had an efficacy of 74% 28 days after the first dose. Lastly, seven of eight studies found significantly increased cycle threshold, suggestive of lower viral load, in PfBnT or AZ vaccinated individuals compared with those who were unvaccinated.

**Conclusion:** The AZ and PfBnT vaccines may prevent household transmission of COVID-19 after 14 days of vaccination. More studies have found the vaccines to significantly reduce the risk of asymptomatic infection and significantly increase cycle threshold, suggestive of lower viral load. Further research is needed to evaluate post-vaccination infectivity and transmission of both the wild type COVID-19 virus and the variants of concern from other jurisdictions.

## Background

Coronavirus disease (COVID-19) is caused by severe acute respiratory syndrome coronavirus 2 (SARS-CoV-2). As of May 2021, there have been more than 164,000,000 confirmed cases of COVID-19, which have resulted in more than 3,400,000 confirmed deaths worldwide.^23^ Since the start of the pandemic, several clinical trials have been underway to examine the safety and effectiveness of different vaccines to prevent COVID-19. Many of these have found the vaccines to be generally effective against symptomatic COVID-19 infection, with an average efficacy of 85% (95% CI: 71 - 93%) after a full course of vaccination.^24^

People who have started or finished the COVID19 vaccine series have been documented to have detectable SARSCoV2 by RT-PCR at various time points after vaccination,^4^ although demonstration of cultivatable virus and definitive evidence of transmission post vaccination has not been assessed. It is not yet clear whether the current COVID-19 vaccines are as effective at reducing transmission as they are at reducing disease. Moreover, evaluating the ability of vaccinated individuals to transmit the virus after infection is challenging. Therefore, virologic surrogates of possible transmissibility may be a helpful way around this challenge.

Monoclonal antibody studies may provide useful insights into the pathophysiologic plausibility of vaccine induced transmission reduction, since they have been shown to result in circulating neutralizing antibody, with a significant decrease in quantitative viral load.^25^ In one study, following quantitative reverse-transcriptase–polymerase-chain-reaction (RT-PCR) testing of nasopharyngeal swabs, an antibody cocktail was found to significantly reduce viral load compared with placebo.^25^ The time-weighted average change in viral load in the first 7 days was −0.56 log10 copies per milliliter (95% CI, −1.02 to −0.11) among those who were serum antibody–negative at baseline.^25^ Another study reported an elimination of more than 99.97% of viral RNA on day 11 after monoclonal antibody treatment.^26^

There is evolving data around the frequency of asymptomatic COVID-19 and if the viral load, and therefore infectiousness, is lower among people who develop COVID-19 post-vaccination compared with those who have not been vaccinated. Viral presence is an imperfect proxy of transmissibility although the quantity of virus present does appear to influence risk, as studies document transmission risk is higher with a higher viral load or lower Ct value.^27, 28^ Marks et al. found index viral load to be a major driver of transmission in a Spanish cohort,^28^ with only 32% of index cases responsible for transmission, and an attack rate of 12% in contacts of index cases with a viral load <10^6^ and 25% in contacts of index cases with a viral load of 10^10^. Similarly, Bjorkman et al. found that higher viral load increased SARS-CoV-2 transmission between asymptomatic residence hall roommates.^29^ The index cases who transmitted infection had an average viral load 6.5 log higher than those who did not. Transmission from asymptomatic students to roommates occurred in 20% of rooms with an infected student, with a lower mean Ct (E gene) of 26.2 in transmission index cases versus 28.9, (median 26.11 in transmission index cases versus 29.32).

However, the risks related to viral presence by RT-PCR may be modulated by individual’s immune status, as viral persistence after natural infection has been observed in individuals with neutralizing antibody responses after natural infection, without transmission to close contacts.^30^

Although asymptomatic and especially pre-symptomatic transmission of SARSCoV-2 has been well documented, existing studies suggest that transmission risk is lower from asymptomatic individuals than symptomatic individuals.^31^

The evidence for the transmissibility and transmission of COVID-19 infections in vaccinated individuals is rapidly evolving; therefore, the objective of this rapid review was to identify comparative observational studies and randomized controlled trials (RCTs) evaluating the effectiveness or efficacy of COVID-19 vaccination in reducing infection transmission, asymptomatic viral carriage and other proxies of possible transmission, such as cycle threshold (Ct) values and viral load. This is an update of a previous report with a literature search that ended 11 March 2021.^1^

## Methods

An experienced medical information specialist developed and tested the search strategies through an iterative process in consultation with the review team. The MEDLINE strategy was peer reviewed by another senior information specialist prior to execution using the PRESS Checklist.^32^

Using the multifile option and deduplication tool available on the OVID platform, we searched Ovid MEDLINE®, including Epub Ahead of Print, In-Process & Other Non-Indexed Citations, Embase, and EBM Reviews - Cochrane Central Register of Controlled Trials. We also searched for primary studies on the Living Overviews of Evidence (L-OVE) platform. We performed all searches on May 4, 2021.

The strategies utilized a combination of controlled vocabulary (e.g., “COVID-19 Vaccines”, “COVID-19/tm [Transmission]”, “Disease Transmission, Infectious”) and keywords (e.g., “mRNA vaccine”, “unvaccinated”, “infectiousness”). Vocabulary and syntax were adjusted across the databases. The search strategies are in Appendix 1. No language or date limits were applied. Results were downloaded and deduplicated using EndNote version 9.3.3 (Clarivate Analytics) and uploaded to word.

A grey literature search was also conducted, including: Clinicaltrials.gov, McMaster Health Forum (CoVID-END), MedRxiv, Google, regulatory submissions, and websites of the Center for Disease Control and Prevention (CDC) and World Health Organization (WHO). This search was limited to studies conducted in 2020 and 2021 and current to May 4th, 2021. There were no language limitations.

A screening form based on the eligibility criteria was prepared. Citations identified as potentially relevant from the literature search were screened by a reviewer, and subsequently read in full text by two reviewers and assessed for eligibility based on the criteria outlined below (Table 1). Discrepancies were resolved by discussion or by a third reviewer. Reference lists of included studies were hand searched to ensure all relevant literature is captured.

**Table 1.** Criteria for Inclusion

|  |  |
| --- | --- |
| Population | Persons who had received COVID-19 vaccination irrespective of age, sex or gender. Animal studies were also included. |
| Intervention | COVID-19 vaccination |
| Comparator | Non-vaccinated persons. |
| Outcome | Include, but are not limited to, Ct values, viral load, asymptomatic laboratory confirmed cases by RT-PCR post-vaccination and the number of persons who are infected by someone who has COVID-19 and has had the vaccine. Studies evaluating the transmissibility or infectivity of COVID-19 among vaccinated individuals were included. |
| Study Design | Comparative observational studies and RCTs evaluating the efficacy and effectiveness of COVID-19 vaccination in the prevention of asymptomatic viral infections as a proxy of a possible transmission will be included. Studies eligible for inclusion must have a control group of unvaccinated people. |

A standardized data extraction sheet was used to extract the year of publication, country, study design, patient characteristics including sex, gender and age, variants of COVID-19, whether infection was symptomatic or asymptomatic, and all the reported outcomes. All reviewers completed a calibration exercise whereby data from two sample studies were extracted by all four reviewers and areas of disagreement were discussed. Data were extracted by one reviewer and verified by another reviewer.

Quality assessment was conducted based on study design: Cochrane risk of bias for non-randomized studies (ROBINS-I) for non-randomized studies,^33^ Cochrane Risk of Bias (version 5.1.0) for human-subject RCTs,^34^ and the Systematic Review Centre for Laboratory animal Experimentation’s (SYRCLE) risk of bias for animal studies.^35^ Quality assessment was conducted by one reviewer and verified by a second reviewer.

## Results

### Study Characteristics

This updated search yielded 4993 unique citations, 4749 of which were excluded after abstract review (Figure 1). Fifty studies identified from the database search proceeded to full-text review. An additional 82 studies identified through grey literature search were also reviewed. In total, ninety-nine studies were excluded for the following reasons: outcomes not of interest (n=54), study design not of interest (n=10), comparator not of interest (n=4), intervention not of interest (n=1) and duplicate (n=30).

**Figure 1:**
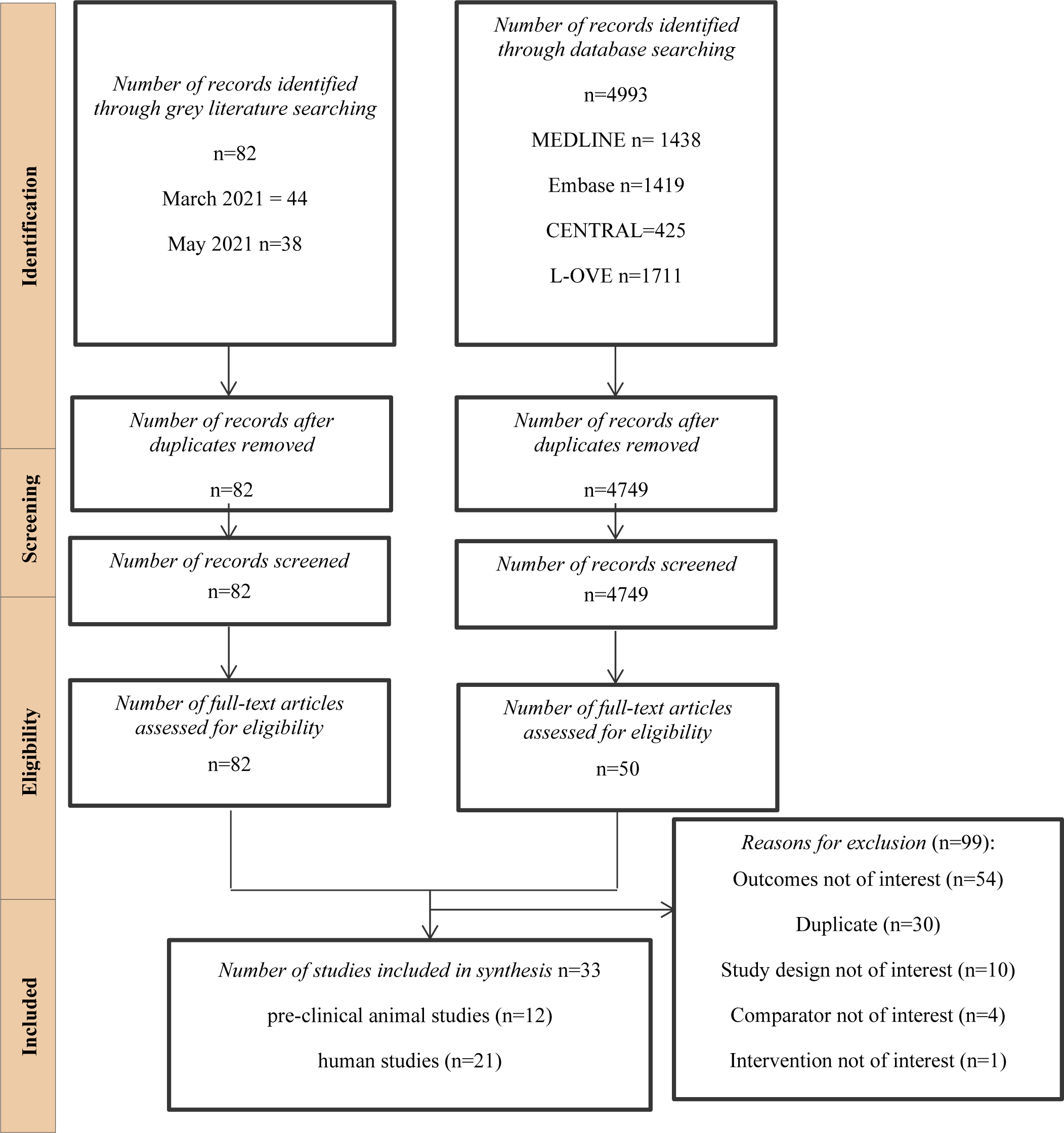
Flowchart of Included Studies

A total of 33 studies were included in this review (Figure 1). Twenty-one were human studies (Table 2 and Table 3),^2–17, 19, 29, 36–38^ and 12 were preclinical animal studies, with viral challenge 1-17 weeks post vaccination (Table 4).^20–22, 39–47^ In the previous report, a targeted literature search was conducted and 17 studies included. Thus, an additional 16 studies have been included since the last update on 11 March 2011, including two studies evaluating household transmission. Five of the human studies were randomized controlled trials,^4–6, 18, 19^ eight were prospective cohort studies,^8, 11^ seven were retrospective cohort studies,^2, 10, 12, 13, 36–38^ and one was a case control study.^15^

**Table 2:** Characteristics of Included RCTs

| Author/Country/Design | Trial Information | Participant Inclusion/Exclusion Criteria | Vaccine Information | Efficacy/Effectiveness Outcomes |
| --- | --- | --- | --- | --- |
| **Author:** Voysey 2021<sup>5</sup><br>**County:** UK, Brazil, S.Africa<br>**Date of Recruitment:** May-Nov 2020<br>**Trial Phase:** 2/3<br>**Design:** Single Blind RCT<br>**Funding:** UK Research and Innovation, National Institutes for Health Research (NIHR), Coalition for Epidemic Preparedness Innovations, Bill & Melinda Gates Foundation, Lemann Foundation, Rede D'Or, Brava and Telles Foundation, NIHR Oxford Biomedical Research Centre, Thames Valley and South Midland's NIHR Clinical Research Network, and AstraZeneca. | **Age:** NR<br>**%Female:** Varied<br>**Type of comparator:** Meningococcal vaccine<br>**Sample Size Vaccine:** Varied<br>**Sample Size Control:** Varied<br>**Total Sample:** Varied<br>**VOC:** NR | Healthy volunteers aged over 18; at risk of virus, stable pre-existing conditions | **Vaccine:** ChAdOx1 nCoV-19<br>**Manufacturer:** AstraZeneca<br>**Dose:** Low or Standard Doses<br>**Number of Doses:** 2 | • Symptomatic Infection<br>• Severe Cases<br>• Asymptomatic infection (weekly self- administered nose and throat swab for NAAT testing from 1 week after first vaccination using kits provided by the UK Department of Health and Social Care) |
| **Author:** Voysey, 2021<sup>4</sup><br>**County:** UK, Brazil, S.Africa<br>**Date of Recruitment:** May-Dec 2020<br>**Trial Phase:** 1/2/3<br>**Design:** Single Blind RCT<br>**Funding:** UKRI, NIHR, CEPI, the Bill & Melinda Gates Foundation, the Lemann Foundation, Rede D'OR, the Brava and | **Age:** NR<br>**%Female:** NR<br>**Type of comparator:** Meningococcal vaccine<br>**Sample Vaccine:** 8567<br>**Sample Control:** 8580<br>**Total Sample:** 17177 | NR | **Vaccine:** ChAdOx1 nCoV-19<br>**Manufacturer:** AstraZeneca<br>**Dose:** Low or Standard Doses<br>**Number of Doses:** 2 | • Symptomatic Infection<br>• Severe Cases<br>• Asymptomatic infection (measured by means of weekly self-administered nose and throat swabs using kits provided by the Department of Health and Social Care) |
| Telles Foundation, NIHR Oxford Biomedical Research Centre, Thames Valley and South Midland's NIHR Clinical Research Network, and Astra Zeneca | VOC: NR |  |  |  |
| **Author:** Emary 2021<sup>6</sup><br>**County:** UK<br>**Date of Recruitment:** Oct-Jan 2021<br>**Trial Phase:** 2/3<br>**Design:** RCT<br>**Funding:** UK Research and Innovation, National Institutes for Health Research (NIHR), Coalition for Epidemic Preparedness Innovations, NIHR Oxford Biomedical Research Centre, Thames Valley and South Midlands NIHR Clinical Research Network, and AstraZeneca. | **Age:** NR<br>**%Female:** NR<br>**Type of comparator:** Meningococcal vaccine<br>**Sample Vaccine:** 4236<br>**Sample Control:** 4270<br>**Total Sample:** 8506<br>**VOC:** B.1.1.7, Other | Aged 18 and over; high-exposure populations eligible for vaccination under the government National Health Service coronavirus vaccine programme. | **Vaccine:** ChAdOx1 nCoV-19<br>**Manufacturer:** AstraZeneca<br>**Dose:** Low or Standard Doses<br>**Number of Doses:** 2 | Symptomatic Infection<br>Ct Values (weekly swabs processed. The minimum Ct value across the N and ORF1ab genes from each PCR test was computed)<br>Asymptomatic Unknown infection (upper airway swabs every week during the trial. Cases were excluded if they occurred before 15 days post the second dose of vaccine or occurred in participants who were not seronegative on a SARS-CoV-2 N protein assay at baseline) |
| **Author:** Janssen Biotech, 2021<sup>19</sup> (Regulatory Submission)<br>**County:** Argentina, Brazil, Chile, Colombia, Mexico, Peru, South Africa, and the United States<br>**Date of Recruitment:** Sept 2020-Jan 2021<br>**Trial Phase:** 3 | **Age:** 51.1 (15.0)<br>**%Female:** 44.5<br>**Comparator:** Placebo<br>**Sample Vaccine:** 19514<br>**Sample Control:** 19544 | Adults 18+ with or without comorbidities. | **Vaccine:** Ad26.COV2.S<br>**Manufacturer:** Janssen Biotech<br>**Dose:** NR<br>**Number of Doses:** 1 | Severe cases<br>Moderate to Severe infections<br>Asymptomatic infection (No symptoms on the day preceding, the day of, or any time after the positive PCR test AND has a SARS-CoV-2 positive RT-PCR test result OR develops a |
| **Design:** Double Blind RCT<br>**Funding:** Janssen Biotech | **Total Sample:** 39058<br>VOC:NR |  |  | positive serology based on a SARS-CoV-2 N-specific immunoglobulin assay (Elecsys®, Roche) during the study. SARS CoV-2 seropositivity by non-S protein was assessed at Day 1 (pre-vaccination), Day 29 (28 days post-vaccination), and Day 71) |
| **Author:** Baden, 2021<sup>18</sup><br>**County:** USA<br>**Date of Recruitment:** Jul-Nov, 2020<br>**Trial Phase:** 3<br>**Design:** Observer Blinded RCT<br>**Funding:** Biomedical Advanced Research and Development Authority and the National Institute of Allergy and Infectious Diseases | **Age:** 51.4<br>**%Female:** 47.3<br>**Comparator:** saline<br>**Sample Vaccine:** 14550<br>**Sample Control:** 14598<br>**Total Sample:** 29148 | **Include:** Eligible participants were persons 18 years of age or older with no known history of SARS-CoV-2 infection and with locations or circumstances that put them at an appreciable risk of SARSCoV-2 infection, a high risk of severe COVID-19, or both.<br>**Exclude:** Pregnant women and children | **Vaccine:** mRNA-1273<br>**Manufacturer:** Moderna<br>**Dose:** 100mcg<br>**Number of Doses:** 2 | • Symptomatic infection<br>• Severe cases<br>• Any Positive PCR<br>• Asymptomatic infection (Surveillance swab at the second dose visit) |

IQR: interquartile range, NR: Not Reported, PCR: Polymerase Chain Reaction, RCT: randomized controlled trial, VOC: Variant of Concern, Studies are peer reviewed publications except otherwise stated.

**Table 3:** Characteristics of Observational Studies

| Author/Country/Design | Trial Information | Participant Inclusion/Exclusion Criteria | Vaccine Information | Effectiveness Outcomes |
| --- | --- | --- | --- | --- |
| **Author:** Hall, 2021<sup>11</sup><br>**County:** UK<br>**Date of Recruitment:** Dec 2020-Feb, 2021<br>**Trial Phase:** Post Approval<br>**Design:** Prospective Cohort<br>**Funding:** Public Health England and the Department of Health and Social Care; NIHR | **Age:** NR<br>**%Female:** 84<br>**Type of comparator:** Unvaccinated<br>**Sample Vaccine:** NR<br>**Sample Control:** NR<br>**Total Sample:** NR<br>**VOC:** B.1.1.7 | Health care workers at hospital, who could provide informed consent and anticipated remaining engaged in follow-up for 12 months. | **Vaccine:** BNT162b2<br>**Manufacturer:** Pfizer BioNTech<br>**Dose:** NR<br>**Number of Doses:** 1 or 2 | Symptomatic Infection<br>Asymptomatic infection (fortnightly asymptomatic PCR testing (anterior nasal swabs or combined nose and oropharyngeal swabs) and monthly antibody testing)<br>Any positive PCR |
| **Author:** Amit, 2021<sup>12</sup><br>**County:** Israel<br>**Date of Recruitment:** Dec 2020-Jan 2021<br>**Trial Phase:** Post Approval<br>**Design:** Retrospective Cohort<br>**Funding:** NR | **Age:** NR<br>**%Female:** NR<br>**Comparator:** Unvaccinated<br>**Sample Vaccine:** NR<br>**Sample Control:** NR<br>**Total Sample:** NR<br>**VOC:** NR | NR | **Vaccine:** BNT162b2<br>**Manufacturer:** Pfizer BioNTech<br>**Dose:** 1 or 2<br>**Number of Doses:** 2 | Symptomatic Infection<br>Any positive PCR |
| **Author:** Dagan, 2021<sup>8</sup><br>**County:** Israel<br>**Date of Recruitment:** Dec 2020-Feb, 2021 | **Age:** Unvaccinated: 45 (IQR:35–62), vaccinated: 45 (35–62)<br>**%Female:** 50 | **Include:** 16 years or older, not having a previously documented positive SARS-CoV-2 PCR test, and being a member of the | **Vaccine:** BNT162b2<br>**Manufacturer:** Pfizer BioNTech | Symptomatic Infection<br>Severe Cases |
| **Trial Phase:** Post Approval<br>**Design:** Prospective Cohort<br>**Funding:** NR | **Comparator:** Unvaccinated<br>**Sample Vaccine:** 596618<br>**Sample Control:** 596618<br>**Total Sample:** 1193236<br>**VOC:** B.1.1.7 | health care organization during the previous 12 months.<br>**Exclude:** probability of exposure or the outcomes is high and controlling for the high variability is not feasible. | **Dose:** NR<br>**Number of Doses:**2 | Asymptomatic infection (testing protocol not defined, however SARS-CoV-2 infection without documented symptoms used as proxy) |
| **Author:** Levine-Tiefenbrun, 2021<sup>36</sup><br>**County:** Israel<br>**Date of Recruitment:** Dec 2020-Jan, 2021<br>**Trial Phase:** Post Approval<br>**Design:** Retrospective Cohort<br>**Funding:** NR | **Age:** NR<br>**%Female:** NR<br>**Comparator:** Unvaccinated<br>**Sample Vaccine:** Varied<br>**Sample Control:** Varied<br>**Total Sample:** Varied<br>**VOC:** NR | **Include:** All positive post-vaccination samples;<br>**Exclude:** Patients who had a positive sample prior to vaccination; patients age 90 and above | **Vaccine:** BNT162b2<br>**Manufacturer:** Pfizer BioNTech<br>**Dose:** NR<br>**Number of Doses:**1 | Ct values |
| **Author:** Jones, 2021<sup>13</sup><br>**Country:** UK<br>**Date of Recruitment:** Jan 18-31, 2021<br>**Trial Phase:** Post Approval<br>**Design:** Retrospective Cohort<br>**Funding:** Wellcome Senior Clinical Research Fellowship to MPW | **Age:** NR<br>**%Female:** NR<br>**Comparator:** Unvaccinated<br>**Sample Vaccine:** 3535<br>**Sample Control:** 3252 | **Include:** vaccinated and unvaccinated Health Care Workers<br>**Exclude:** NR | **Vaccine:** BNT162b2<br>**Manufacturer:** Pfizer BioNTech<br>**Number of Doses:**1 | Any positive PCR<br>Ct values<br>Asymptomatic (weekly Screening) |
| (108070/Z/15/Z), a Wellcome Principal Research Fellowship to PJL (210688/Z/18/Z), and an MRC Clinician Scientist Fellowship (MR/P008801/1) and NHSBT workpackage (WPA15-02) to NJM. Funding was also received from Addenbrooke's Charitable Trust and the Cambridge Biomedical Research Centre. | **Total Sample:** Varied<br>**VOC:** B.1.1.7 |  |  |  |
| **Author:** Tande, 2021<sup>10</sup><br>**Country:** USA<br>**Date of recruitment:** December, 2020 to February, 2021<br>**Trial Phase:** Post approval<br>**Design:** Retrospective Cohort<br>**Funding:** Internal funding at the Mayo Clinic | **Age:** 54.2 (19.7)<br>**%Female:** 52.5<br>**Comparator:** Unvaccinated<br>**Sample vaccine:** 3006<br>**Sample control:** 45,327<br>**Total sample:**<br>**VOC:** NR | **Include:** 18 or more years old, underwent preprocedural/presurgical testing within 48-72 hours of procedure<br>**Exclude:** Patients tested due to symptoms or a known exposure were tested using an alternative ordering process | **Vaccine:** BNT162b2 or mRNA-1273<br>**Manufacturers:** Pfizer BioNTech or Moderna<br>**Number of Doses:** 1 or 2 | PCR+ among Asymptomatic (consecutive preprocedural molecular screening tests) |
| **Author:** Pfizer BioNTech<sup>38</sup> (Press Release)<br>**Country:** Israel<br>**Date of recruitment:** January 17 through March 6, 2021<br>**Trial Phase:** Post approval<br>**Design:** Retrospective cohort | **Age:** NR<br>**%Female:** NR<br>**Sample vaccine:** NR<br>**Sample control:** NR<br>**Total sample:** NR | NR | **Vaccine:** BNT162b2<br>**Manufacturers:** Pfizer BioNTech | Symptomatic<br>Asymptomatic (Surveillance method unclear) |
| **Author:** McEllistrem, 2021<sup>37</sup> | **Age:** NR | **Include:** | **Vaccine:** BNT162b2 | Ct values |
| **Country:** USA<br>**Date of recruitment:** December 8, 2020–February 2, 2021<br>**Trial Phase:** Post approval<br>**Design:** Retrospective Cohort<br>**Funding:** None | **%Female:** NR<br>**Comparator:** Unvaccinated<br>**Sample vaccine:** 5<br>**Sample control:** 5<br>**Total sample:** 10<br>**VOC:** NR | A negative baseline nasopharyngeal reverse transcription polymerase chain reaction test (RT-PCR, Palo Alto VA, CA) for SARS-CoV-2 on 12/2/20. | **Manufacturers:** Pfizer BioNTech<br>**Number of Doses:** 1 | • Viral load<br>• Asymptomatic (surveillance nares testing for SARS-CoV-2 with the BD Veritor antigen every 2-5 days) |
| **Author:** Shah, 2021<sup>2</sup> (Pre-print)<br>**Country:** UK<br>**Date of recruitment:** December 8, 2020 – March 3, 2021<br>**Trial Phase:** Post approval<br>**Design:** Retrospective Cohort<br>**Funding:** British Heart Foundation through an intermediate clinical research fellowship (FS/19/17/34172); Wellcome Trust intermediate clinical fellowship and Beit fellowship (201492/Z/16/Z) | **Age:** 44.4(11.4)<br>**%Female:** 78.7<br>**Comparator:** Unvaccinated<br>**Sample vaccine:** 109,074<br>**Sample control:** 144,525<br>**Total sample:**<br>**VOC:** NR | **Include:** Healthcare workers were included if they were employed by the National Health Service (NHS) in Scotland on or before the 1st of March 2020 (the first positive reported case of COVID-19 in Scotland) and still employed by the NHS on the 1st of November 2020; healthcare worker cohort was restricted to the working-age population (18-65 years of age). The household member cohort included all ages but was restricted to households with no more than one healthcare worker (4% of healthcare workers lived in multiple healthcare worker households) | **Vaccine:** BNT162b2 or ChAdOx1 nCoV-19<br>**Manufacturers:** Pfizer BioNTech or Oxford AstraZeneca<br>**Number of Doses:** 1 | • Transmission to contact |
| **Author:** Bouton, 2021<sup>15</sup> (Pre-print)<br>**Country:** USA<br>**Date of recruitment:** December 9, 2020-February 23, 2021<br>**Trial Phase:** Post approval<br>**Design:** Case Control<br>**Funding:** | **Age:** 40(13)<br>**%Female:** NR<br>**Comparator:** Unvaccinated<br>**Sample vaccine:** 96<br>**Sample control:** 329<br>**Total sample:** 425<br>**VOC:** NR | **Include:** HCWs had been vaccinated prior to the vaccine initiative and were included in analyses. HCW who received a vaccination following their positive SARS-CoV-2 RT-PCR were included in the unvaccinated group. | **Vaccine:** BNT162b2 or mRNA-1273<br>**Manufacturers:** Pfizer BioNTech or Moderna<br>**Number of Doses:** 1 | Asymptomatic (Asymptomatic testing is available to HCWs for workplace exposures, following out-of-state travel, and per request)<br>All PCR-positive (symptomatic and asymptomatic) |
| **Author:** Regev-Yochay, 2021<sup>14</sup> (Pre-print)<br>**Country:** Israel<br>**Date of recruitment:** December 19, 2020 – March 14, 2021<br>**Trial Phase:** Post approval<br>**Design:** Cohort<br>**Funding:** Sheba Medical Center, Israel | **Age:** NR<br>**%Female:** NR<br>**Comparator:** Unvaccinated<br>**Sample vaccine:**<br>**Sample control:**<br>**Total sample:** 3578<br>**VOC:** NR | **Include:** HCW at Sheba Medical Center (Israel) | **Vaccine:** BNT162b2 or mRNA-1273<br>**Manufacturers:** Pfizer BioNTech or Moderna<br>**Number of Doses:** 1 or 2 | Asymptomatic (Symptomatic or exposed to confirmed case)<br>Symptomatic<br>Severe cases |
| **Author:** Lumley, 2021<sup>17</sup> (Pre-print)<br>**Country:** England<br>**Date of recruitment:** Through to February 28, 2021 | **Age:** 39 (IQR:30-50)<br>**%Female:** 74.0<br>**Comparator:** Unvaccinated seronegative | **Include:** Only those who participated in asymptomatic screening, symptomatic testing or vaccination from 01-September-2020 onwards were included. All staff working for | **Vaccine:** BNT162b2 or ChAdOx1 nCoV-19<br>**Manufacturers:** Pfizer BioNTech or Oxford AstraZeneca | Ct values<br>Symptomatic<br>Asymptomatic Asymptomatic (voluntary nasal and oropharyngeal swab) |
| **Trial Phase:** Post approval<br>**Design:** Longitudinal Cohort<br>**Funding:** Supported by the UK Government's Department of Health and Social Care. Also supported by the National Institute for Health Research Health Protection Research Unit (NIHR HPRU) in Healthcare Associated Infections and Antimicrobial Resistance at Oxford University in partnership with Public Health England (PHE) (NIHR200915), the NIHR Biomedical Research Centre, Oxford, and benefactions from the Huo Family Foundation and Andrew Spokes. | **Sample vaccine:** NR<br>**Sample control:** NR<br>**Total sample:** 13,109<br>**VOC:** B.1.1.7 | the hospitals were eligible to participate. | **Number of Doses:** 1 or 2 | PCR testing every two weeks and serological testing every two months) |
| **Author:** Pritchard, 2021<sup>7</sup> (Pre-print)<br>**Country:** UK<br>**Date of recruitment:** December 1, 2020 – April 3, 2021<br>**Trial Phase:** Post approval<br>**Design:** Prospective Cohort<br>**Funding:** Department of Health and Social Care with in-kind support from the Welsh Government, the Department of Health on behalf of the Northern Ireland Government and the Scottish Government. | **Age:** NR<br>**%Female:** NR<br>**Comparator:** Unvaccinated<br>**Sample vaccine:**<br>**Sample control:**<br>**Total sample:** 373,402<br>**VOC:** NR | **Include:** This analysis included participants aged 16 years or over (i.e. those who theoretically could have received vaccination), and all visits with positive or negative swab results from 1 December 2020 to 3 April 2021. | **Vaccine:** BNT162b2 or ChAdOx1 nCoV-19<br>**Manufacturers:** Pfizer BioNTech or Oxford AstraZeneca<br>**Number of Doses:** 1 or 2 | Asymptomatic (Weekly nose and throat self-swab for first month, then monthly for 12 months from enrolment)<br>Ct values<br>Symptomatic |
| **Author:** Shrotri, 2021.<sup>16</sup> (Pre-print)<br>**Country:** UK<br>**Date of recruitment:** December 8, 2020 – March 15, 2021<br>**Trial Phase:** Post approval<br>**Design:** Prospective Cohort<br>**Funding:** UK Government Department of Health and Social Care. | **Age:** 86 (IQR: 80-91)<br>**%Female:** 69.6<br>**Comparator:** Unvaccinated<br>**Sample vaccine:**<br>**Sample control:**<br>**Total sample:** 10,412<br>**VOC:** NR | **Include:** At least two PCR test results in total, and $\geq 1$ PCR result during the analysis period. Residents entered the risk period on 8 December 2020 if they had $\geq 1$ valid PCR result on or prior to that date; or, if they had no PCR results before 8 December 2020, on the date of their first negative PCR test. Residents with a positive PCR result $\leq 90$ days before 8 December entered the risk period 90 days after their positive test. | **Vaccine:** BNT162b2 or ChAdOx1 nCoV-19<br>**Manufacturers:** Pfizer BioNTech or Oxford AstraZeneca<br>**Number of Doses:** 1 or 2 | • Ct values<br>• Symptomatic |
| **Author:** Haas, 2021<sup>9</sup><br>**Country:** Israel<br>**Date of recruitment:** Jan 24, 2021–April 3, 2021<br>**Trial Phase:** Post approval<br>**Design:** Prospective Cohort<br>**Funding:** Israel MoH and Pfizer. | **Age:** NR<br>**%Female:** 50.8<br>**Comparator:** Unvaccinated<br>**Sample vaccine:** NR<br>**Sample control:** NR<br>**Total sample:** NR<br>**VOC:** B.1.1.7 | **Include:** unvaccinated and vaccinated individuals aged $\geq 16$ years. | **Vaccine:** BNT162b2<br>**Manufacturers:** Pfizer BioNTech<br>**Number of Doses:** 1 or 2 | • Asymptomatic (routine testing)<br>• Severe cases<br>• Symptomatic |
| **Author:** Harris, 2021<sup>3</sup> (Pre-print)<br>**Country:** UK | **Age:** NR<br>**%Female:** Unvaccinated index case: 47.6%, Index | **Include:** Households with an index case occurring between 4 January 2021 to 28 February 2021, with 14 days observable | **Vaccine:** BNT162b2 or ChAdOx1 nCoV-19 | • Transmission to contact |
| **Date of recruitment:** January 4 – February 28, 2021<br>**Trial Phase:** Post approval<br>**Design:** Prospective Cohort<br>**Funding:** This work was undertaken as part of the core functions of Public Health England in relation to the surveillance of communicable diseases and outbreak response | case vaccinated 21+ day before: 38.3%, Index case vaccinated <21 days before: 40.6%<br>**Comparator:** Unvaccinated<br>**Sample vaccine:**<br>**Sample control:**<br>**Total sample:** 1,018,842<br>**VOC:** NR | follow up for all contacts; households with a single index case age 16+, and no co-primary cases. | **Manufacturers:** Pfizer BioNTech or Oxford AstraZeneca<br>**Number of Doses:** 1 |  |

IQR: interquartile range, NR: Not Reported, PCR: Polymerase Chain Reaction, RCT: randomized controlled trial, VOC: Variant of Concern, NR: Not Reported. Studies are peer-reviewed publications except otherwise stated.

**Table 4:**
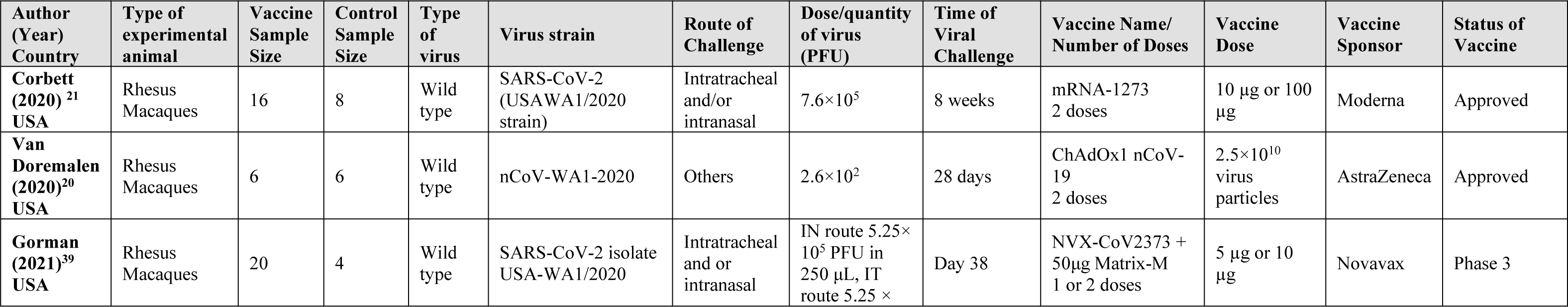
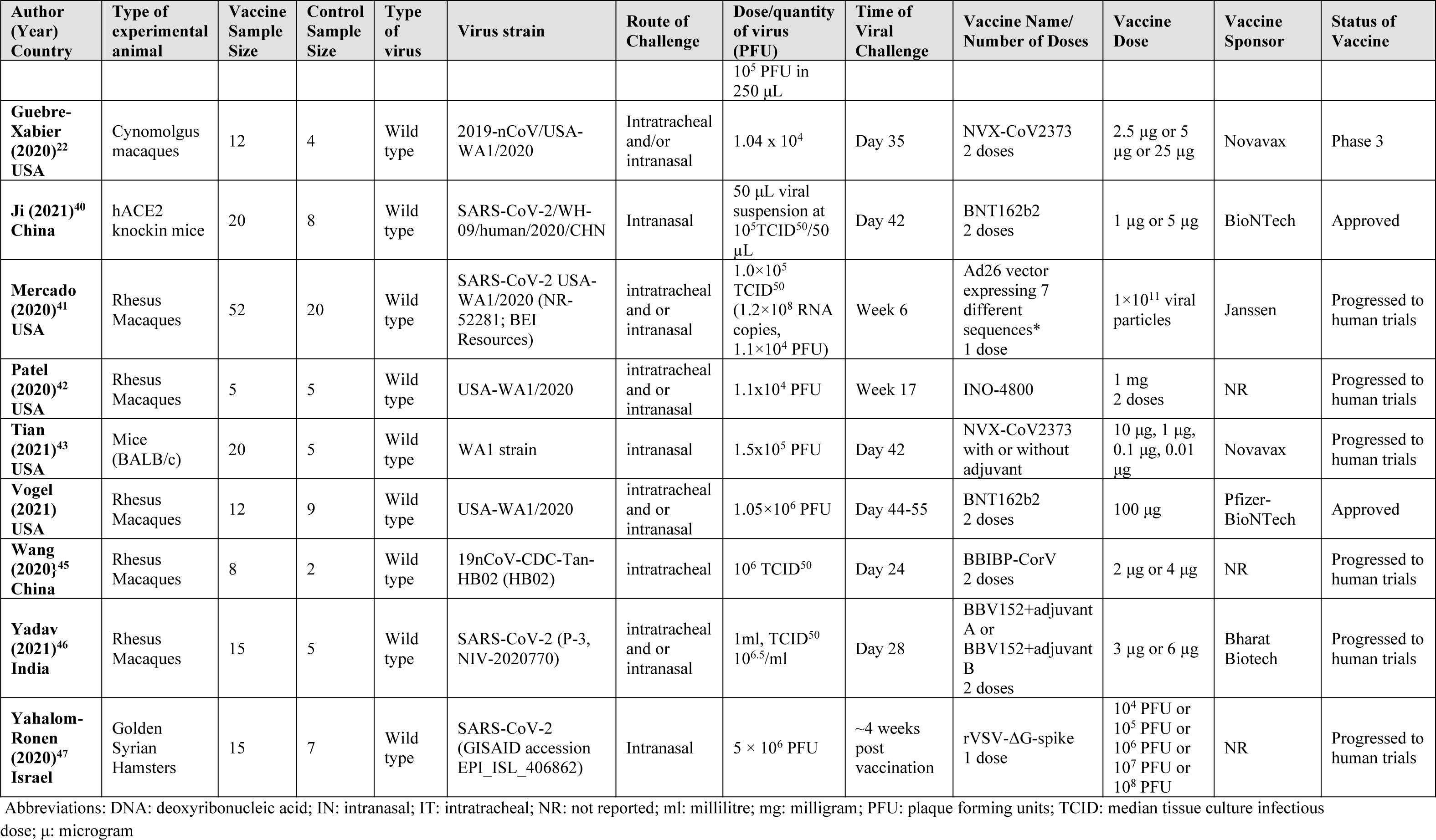
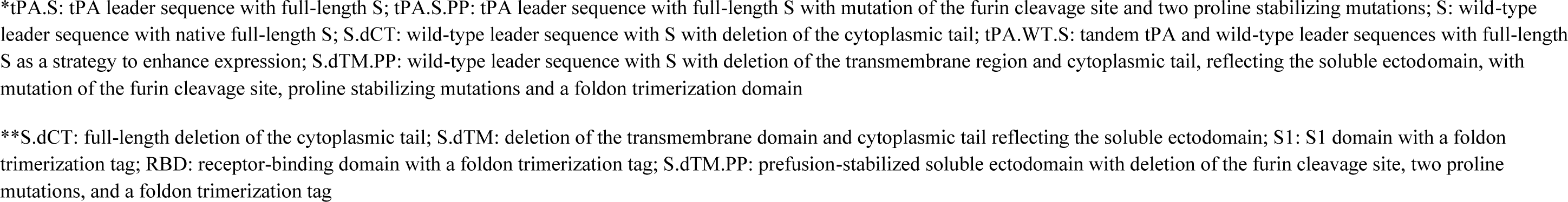
Characteristics of Included Peer-Reviewed Animal Studies

| Author (Year) Country | Type of experimental animal | Vaccine Sample Size | Control Sample Size | Type of virus | Virus strain | Route of Challenge | Dose/quantity of virus (PFU) | Time of Viral Challenge | Vaccine Name/ Number of Doses | Vaccine Dose | Vaccine Sponsor | Status of Vaccine |
| --- | --- | --- | --- | --- | --- | --- | --- | --- | --- | --- | --- | --- |
| **Corbett (2020)**<sup>21</sup> USA | Rhesus Macaques | 16 | 8 | Wild type | SARS-CoV-2 (USAWA1/2020 strain) | Intratracheal and/or intranasal | $7.6 \times 10^5$ | 8 weeks | mRNA-1273 2 doses | 10 µg or 100 µg | Moderna | Approved |
| **Van Doremalen (2020)**<sup>20</sup> USA | Rhesus Macaques | 6 | 6 | Wild type | nCoV-WA1-2020 | Others | $2.6 \times 10^2$ | 28 days | ChAdOx1 nCoV-19 2 doses | $2.5 \times 10^{10}$ virus particles | AstraZeneca | Approved |
| **Gorman (2021)**<sup>39</sup> USA | Rhesus Macaques | 20 | 4 | Wild type | SARS-CoV-2 isolate USA-WA1/2020 | Intratracheal and or intranasal | IN route $5.25 \times 10^5$ PFU in 250 µL, IT route $5.25 \times$ | Day 38 | NVX-CoV2373 + 50µg Matrix-M 1 or 2 doses | 5 µg or 10 µg | Novavax | Phase 3 |
|  |  |  |  |  |  |  | 10<sup>5</sup> PFU in 250 µL |  |  |  |  |  |
| **Guebre-Xabier (2020)<sup>22</sup> USA** | Cynomolgus macaques | 12 | 4 | Wild type | 2019-nCoV/USA-WA1/2020 | Intratracheal and/or intranasal | 1.04 x 10<sup>4</sup> | Day 35 | NVX-CoV2373 2 doses | 2.5 µg or 5 µg or 25 µg | Novavax | Phase 3 |
| **Ji (2021)<sup>40</sup> China** | hACE2 knockin mice | 20 | 8 | Wild type | SARS-CoV-2/WH-09/human/2020/CHN | Intranasal | 50 µL viral suspension at 10<sup>5</sup>TCID<sub>50</sub>/50 µL | Day 42 | BNT162b2 2 doses | 1 µg or 5 µg | BioNTech | Approved |
| **Mercado (2020)<sup>41</sup> USA** | Rhesus Macaques | 52 | 20 | Wild type | SARS-CoV-2 USA-WA1/2020 (NR-52281; BEI Resources) | intratracheal and or intranasal | 1.0×10<sup>5</sup> TCID<sub>50</sub> (1.2×10<sup>8</sup> RNA copies, 1.1×10<sup>4</sup> PFU) | Week 6 | Ad26 vector expressing 7 different sequences* 1 dose | 1×10<sup>11</sup> viral particles | Janssen | Progressed to human trials |
| **Patel (2020)<sup>42</sup> USA** | Rhesus Macaques | 5 | 5 | Wild type | USA-WA1/2020 | intratracheal and or intranasal | 1.1x10<sup>4</sup> PFU | Week 17 | INO-4800 | 1 mg 2 doses | NR | Progressed to human trials |
| **Tian (2021)<sup>43</sup> USA** | Mice (BALB/c) | 20 | 5 | Wild type | WA1 strain | intranasal | 1.5x10<sup>5</sup> PFU | Day 42 | NVX-CoV2373 with or without adjuvant | 10 µg, 1 µg, 0.1 µg, 0.01 µg | Novavax | Progressed to human trials |
| **Vogel (2021) USA** | Rhesus Macaques | 12 | 9 | Wild type | USA-WA1/2020 | intratracheal and or intranasal | 1.05×10<sup>6</sup> PFU | Day 44-55 | BNT162b2 2 doses | 100 µg | Pfizer-BioNTech | Approved |
| **Wang (2020)<sup>45</sup> China** | Rhesus Macaques | 8 | 2 | Wild type | 19nCoV-CDC-Tan-HB02 (HB02) | intratracheal | 10<sup>6</sup> TCID<sub>50</sub> | Day 24 | BBIBP-CorV 2 doses | 2 µg or 4 µg | NR | Progressed to human trials |
| **Yadav (2021)<sup>46</sup> India** | Rhesus Macaques | 15 | 5 | Wild type | SARS-CoV-2 (P-3, NIV-2020770) | intratracheal and or intranasal | 1ml, TCID<sub>50</sub> 10<sup>6.5</sup>/ml | Day 28 | BBV152+adjuvant A or BBV152+adjuvant B 2 doses | 3 µg or 6 µg | Bharat Biotech | Progressed to human trials |
| **Yahalom-Ronen (2020)<sup>47</sup> Israel** | Golden Syrian Hamsters | 15 | 7 | Wild type | SARS-CoV-2 (GISAID accession EPI_ISL_406862) | Intranasal | 5 × 10<sup>6</sup> PFU | ~4 weeks post vaccination | rVSV-ΔG-spike 1 dose | 10<sup>4</sup> PFU or 10<sup>5</sup> PFU or 10<sup>6</sup> PFU or 10<sup>7</sup> PFU or 10<sup>8</sup> PFU | NR | Progressed to human trials |

Abbreviations: DNA: deoxyribonucleic acid; IN: intranasal; IT: intratracheal; NR: not reported; ml: millilitre; mg: milligram; PFU: plaque forming units; TCID: median tissue culture infectious dose; µ: microgram
\*tPA.S: tPA leader sequence with full-length S; tPA.S.PP: tPA leader sequence with full-length S with mutation of the furin cleavage site and two proline stabilizing mutations; S: wild-type leader sequence with native full-length S; S.dCT: wild-type leader sequence with S with deletion of the cytoplasmic tail; tPA.WT.S: tandem tPA and wild-type leader sequences with full-length S as a strategy to enhance expression; S.dTM.PP: wild-type leader sequence with S with deletion of the transmembrane region and cytoplasmic tail, reflecting the soluble ectodomain, with mutation of the furin cleavage site, proline stabilizing mutations and a foldon trimerization domain
\*\*S.dCT: full-length deletion of the cytoplasmic tail; S.dTM: deletion of the transmembrane domain and cytoplasmic tail reflecting the soluble ectodomain; S1: S1 domain with a foldon trimerization tag; RBD: receptor-binding domain with a foldon trimerization tag; S.dTM.PP: prefusion-stabilized soluble ectodomain with deletion of the furin cleavage site, two proline mutations, and a foldon trimerization tag

### Risk of Bias Assessment

The five included RCTs^4–6, 18, 19^ were assessed with the Cochrane Risk of Bias Assessment Tool.^34^ Two studies had some concerns regarding randomization,^6, 19^ two were of low risk^5, 18^ and one study had no sufficient information for assessment.^4^ All but one low risk study^18^ was assessed to have some concerns regarding deviation from intended intervention. Four studies were of some concerns for missing outcome data,^4, 5, 18, 19^ one was assessed to be of high risk of bias.^6^ All the studies were of low risk of bias for the measurement of outcomes. All but one study with some concern ^6^ were of low risk for the selection of reported results. ^4–6, 18, 19^ Overall, four of the RCTs were of some concerns for bias^4, 5, 18, 19^ and one had a high risk of bias^6^ (Table 5).

**Table 5:** Risk of Bias Assessment for RCTs

| Author | Randomization | Deviation from intended intervention | Missing outcome data | Measurement of outcome | Selection of reported results | Overall Bias |
| --- | --- | --- | --- | --- | --- | --- |
| Baden et al.<sup>18</sup> | Low | Low | Some concerns | Low | Low | Some concerns |
| Emary et al.<sup>6</sup> | Some concerns | Some concerns | High | Low | Some concerns | High |
| Janssen Biotech<sup>19</sup> | Some concerns | Some concerns | Some concerns | Low | Low | Some concerns |
| Voysey et al.<sup>5</sup> | Low | Some concerns | Some concerns | Low | Low | Some concerns |
| Voysey et al.<sup>4</sup> | NI | Some concerns | Some concerns | Low | Low | Some concerns |

All studies were published in 2021

The sixteen non-RCTs were assessed using the ROBINS-I tool.^33^ Overall, all but one low risk study^15^ were of moderate risk of bias; while one study did not have enough information for risk assessment^12^ (Table 6). Table 7 describes the results of the SYRCLE risk of bias assessment for the animal studies.^35^ The majority of the studies were assessed as having unclear risks of bias for all domains.

**Table 6:**
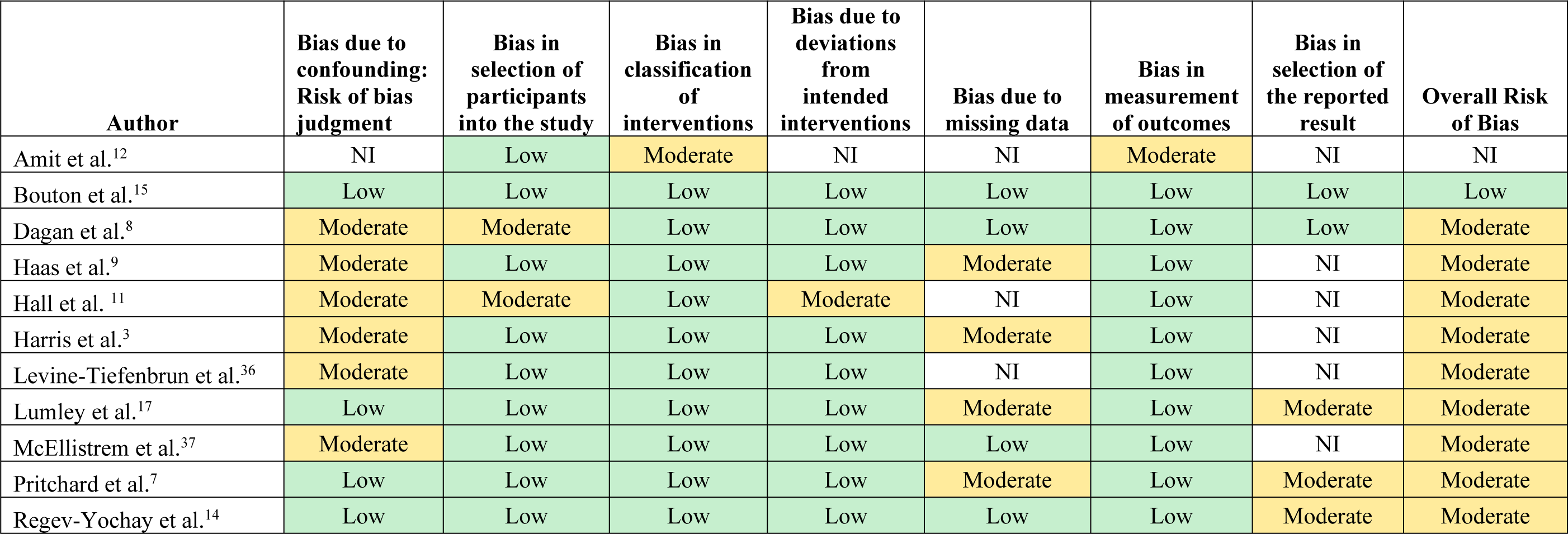
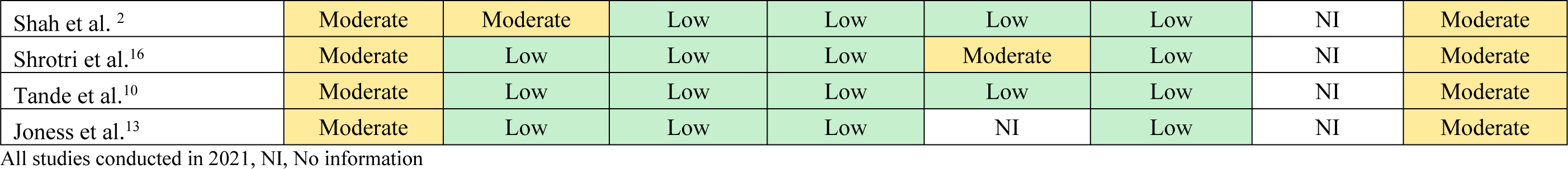
ROBINS-I Risk of Bias for non-RCTs

| Author | Bias due to confounding: Risk of bias judgment | Bias in selection of participants into the study | Bias in classification of interventions | Bias due to deviations from intended interventions | Bias due to missing data | Bias in measurement of outcomes | Bias in selection of the reported result | Overall Risk of Bias |
| --- | --- | --- | --- | --- | --- | --- | --- | --- |
| Amit et al.<sup>12</sup> | NI | Low | Moderate | NI | NI | Moderate | NI | NI |
| Bouton et al.<sup>15</sup> | Low | Low | Low | Low | Low | Low | Low | Low |
| Dagan et al.<sup>8</sup> | Moderate | Moderate | Low | Low | Low | Low | Low | Moderate |
| Haas et al.<sup>9</sup> | Moderate | Low | Low | Low | Moderate | Low | NI | Moderate |
| Hall et al.<sup>11</sup> | Moderate | Moderate | Low | Moderate | NI | Low | NI | Moderate |
| Harris et al.<sup>3</sup> | Moderate | Low | Low | Low | Moderate | Low | NI | Moderate |
| Levine-Tiefenbrun et al.<sup>36</sup> | Moderate | Low | Low | Low | NI | Low | NI | Moderate |
| Lumley et al.<sup>17</sup> | Low | Low | Low | Low | Moderate | Low | Moderate | Moderate |
| McEllistrem et al.<sup>37</sup> | Moderate | Low | Low | Low | Low | Low | NI | Moderate |
| Pritchard et al.<sup>7</sup> | Low | Low | Low | Low | Moderate | Low | Moderate | Moderate |
| Regev-Yochay et al.<sup>14</sup> | Low | Low | Low | Low | Low | Low | Moderate | Moderate |
| Shah et al.<sup>2</sup> | Moderate | Moderate | Low | Low | Low | Low | NI | Moderate |
| Shrotri et al.<sup>16</sup> | Moderate | Low | Low | Low | Moderate | Low | NI | Moderate |
| Tande et al.<sup>10</sup> | Moderate | Low | Low | Low | Low | Low | NI | Moderate |
| Jones et al.<sup>13</sup> | Moderate | Low | Low | Low | NI | Low | NI | Moderate |

All studies conducted in 2021, NI, No information

**Table 7:** SYRCLE Risk of Bias Assessment for Animal Studies

| Author | Allocation Sequence | Similar Baseline | Allocation Concealment | Random Housing of Animals | Blinding of Investigators and Caregivers | Random Selection for Assessment | Blinding of Assessors | Incomplete Outcome Data | Selective Outcome Reporting | Any other possible Bias |
| --- | --- | --- | --- | --- | --- | --- | --- | --- | --- | --- |
| Corbett et al. (2020)<sup>21</sup> | Unclear | Unclear | No | Unclear | Unclear | Unclear | Unclear | Unclear | Unclear | Unclear |
| Van Doremalen et al. (2020)<sup>20</sup> | Yes | Unclear | Unclear | Unclear | Yes | Unclear | Yes | Unclear | Unclear | Unclear |
| Gorman et al. (2021)<sup>39</sup> | Yes | Yes | Yes | Unclear | Yes | Unclear | Yes | Unclear | Unclear | Unclear |
| Guebre-Xabier et al. (2020)<sup>22</sup> | Unclear | Unclear | Unclear | Unclear | Unclear | Unclear | Unclear | Unclear | Unclear | Unclear |
| Ji et al. (2021)<sup>40</sup> | Unclear | Unclear | Unclear | Unclear | Unclear | Unclear | Unclear | Unclear | Unclear | Unclear |
| Mercado et al. (2020)<sup>41</sup> | Unclear | Yes | Unclear | Unclear | Unclear | Unclear | Yes | Unclear | Unclear | Unclear |
| Patel et al. (2020)<sup>42</sup> | Unclear | Unclear | Unclear | Unclear | Unclear | Unclear | Unclear | Unclear | Unclear | Unclear |
| Tian et al. (2021)<sup>43</sup> | Unclear | Unclear | Unclear | Unclear | Unclear | Unclear | Yes | Unclear | Unclear | Unclear |
| Vogel et al. (2021)<sup>44</sup> | No | Unclear | No | Unclear | No | Unclear | No | Unclear | No | Unclear |
| Wang et al. (2020)<sup>45</sup> | Unclear | Unclear | Unclear | Unclear | Unclear | Unclear | Unclear | Unclear | Unclear | Unclear |
| Yadav et al. (2021)<sup>46</sup> | Unclear | Unclear | Unclear | Unclear | Unclear | Unclear | Unclear | Unclear | Unclear | Unclear |
| Yahalom-Ronen et al. (2020)<sup>47</sup> | Unclear | Unclear | Unclear | Unclear | Unclear | Unclear | Unclear | Unclear | Unclear | Unclear |

### Vaccine Efficacy and Effectiveness Against Symptomatic Infection

Several RCTs and observational studies have shown the efficacies and effectiveness of COVID-19 vaccination with ChAdOx1 nCoV-19 (Oxford-AstraZeneca, AZ vaccine), CoronaVac (Sinopharm), BNT162b2 (Pfizer-BionNtech, PfBnT vaccine), Ad26.COV2.S (Janssen, J&J vaccine), mRNA-1273 (Moderna) and NVX-CoV2373 (Novavax) vaccines, against symptomatic COVID-19 infection. Efficacies ranged between 66.1% (95% CI: 55- 74.8) and 94.8% (95% CI 89.8–97.6). These have been compiled and presented in detail in Appendix 2 and Appendix 3. This review however focuses on the transmission of COVID-19 asymptomatic infection and the effect of vaccines on post-infection proxy measures of infectivity such as Ct values and viral load.

### Vaccine Effectiveness against Infection Transmission

Two studies, reported on the effectiveness of both PfBNT and AZ vaccines against disease transmission.

Shah et al. in a retrospective study of 194,362 household members of 144,525 healthcare workers, who had received at least one dose of the PfBnT or AZ, found that from the 14^th^ post-vaccination day onwards, vaccinating a co-habiting healthcare worker was associated with a significantly reduced risk of documented COVID-19 among household members (rate per 100 person-years: 9.40 versus 5.93; HR: 0.70, (95% CI: 0.63-0.78)).^2^ The risk of hospitalization was also significantly lower among household contacts of vaccinated HCWs (rate per 100 person-years: 0.51 versus 0.31; HR: 0.77, (95% CI: 0.53-1.10)).^2^ Following a second dose, the risks of infection and hospitalization involving a household member were significantly lower, rate per 100 person-years of 9.40 versus 2.98, HR: 0.46 (95% CI: 0.30-0.70) and 0.51 versus 0.22 per 100 person-years, HR: 0.68 (95% CI: 0.17-2.83), respectively).^2^ The baseline serology and PCR of household contacts were not reported (Table 8).

**Table 8:**
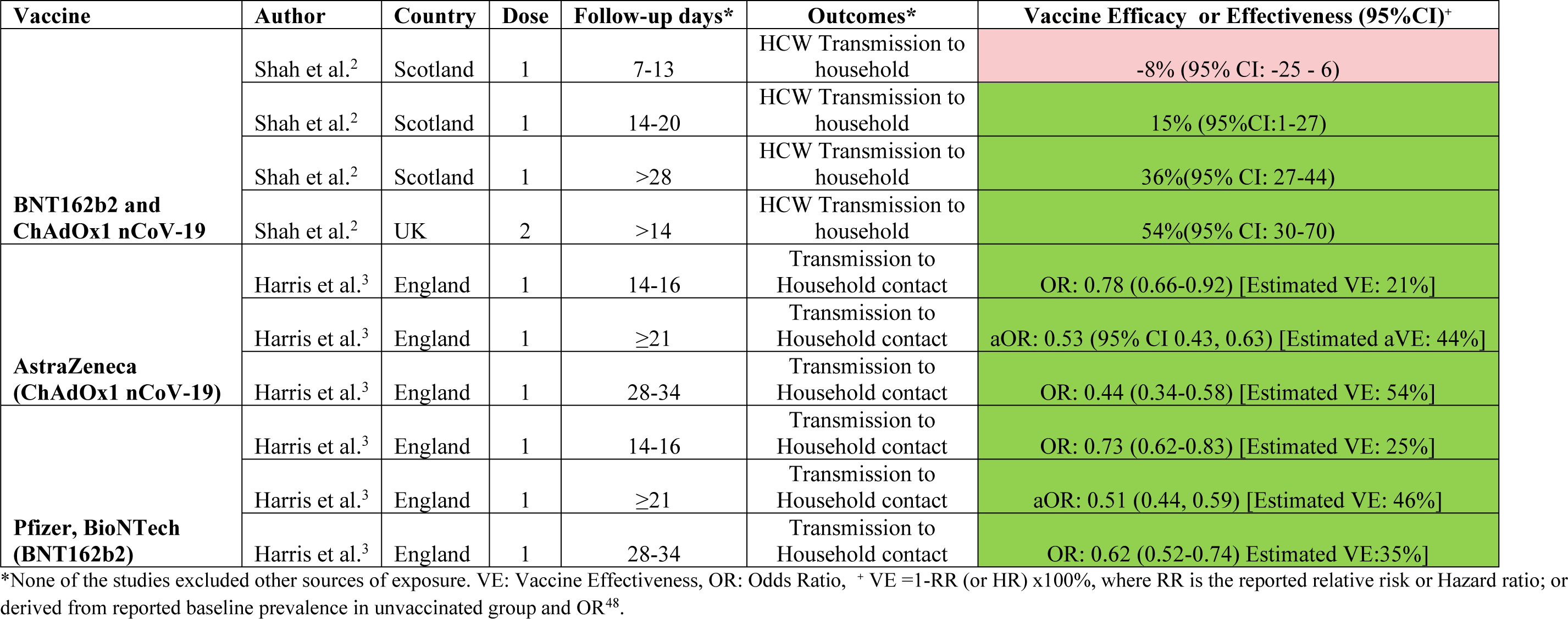
Observational Studies of Vaccine Effectiveness Against Transmission to Household Contacts (Baseline Serology Unknown)

| Vaccine | Author | Country | Dose | Follow-up days* | Outcomes* | Vaccine Efficacy or Effectiveness (95%CI)<sup>+</sup> |
| --- | --- | --- | --- | --- | --- | --- |
| **BNT162b2 and ChAdOx1 nCoV-19** | Shah et al.<sup>2</sup> | Scotland | 1 | 7-13 | HCW Transmission to household | -8% (95% CI: -25 - 6) |
|  | Shah et al.<sup>2</sup> | Scotland | 1 | 14-20 | HCW Transmission to household | 15% (95%CI:1-27) |
|  | Shah et al.<sup>2</sup> | Scotland | 1 | >28 | HCW Transmission to household | 36%(95% CI: 27-44) |
|  | Shah et al.<sup>2</sup> | UK | 2 | >14 | HCW Transmission to household | 54%(95% CI: 30-70) |
| **AstraZeneca (ChAdOx1 nCoV-19)** | Harris et al.<sup>3</sup> | England | 1 | 14-16 | Transmission to Household contact | OR: 0.78 (0.66-0.92) [Estimated VE: 21%] |
|  | Harris et al.<sup>3</sup> | England | 1 | ≥21 | Transmission to Household contact | aOR: 0.53 (95% CI 0.43, 0.63) [Estimated aVE: 44%] |
|  | Harris et al.<sup>3</sup> | England | 1 | 28-34 | Transmission to Household contact | OR: 0.44 (0.34-0.58) [Estimated VE: 54%] |
| **Pfizer, BioNTech (BNT162b2)** | Harris et al.<sup>3</sup> | England | 1 | 14-16 | Transmission to Household contact | OR: 0.73 (0.62-0.83) [Estimated VE: 25%] |
|  | Harris et al.<sup>3</sup> | England | 1 | ≥21 | Transmission to Household contact | aOR: 0.51 (0.44, 0.59) [Estimated VE: 46%] |
|  | Harris et al.<sup>3</sup> | England | 1 | 28-34 | Transmission to Household contact | OR: 0.62 (0.52-0.74) Estimated VE:35%] |

\*None of the studies excluded other sources of exposure. VE: Vaccine Effectiveness, OR: Odds Ratio, <sup>+</sup> VE =1-RR (or HR) x100%, where RR is the reported relative risk or Hazard ratio; or derived from reported baseline prevalence in unvaccinated group and OR<sup>48</sup>.

In a second study, Harris et al. evaluated the risks of transmission of COVID-19 after one dose of PfBnT and AZ vaccination to unvaccinated household contacts using a retrospective design and a matched case-control method.^3^ In the retrospective cohort analysis, there were 96,898 secondary cases among 960,765 household contacts of unvaccinated individuals (10.1%). There were 196 secondary cases in 3,424 contacts (5.72%) where the index case received AZ vaccine more than 21 days before PCR positivity, and 371 secondary cases in 5,939 contacts (6.25%) where the index case received the PfBnT vaccine. Adjusted odds ratio of transmission were 0.53 (95% CI: 0.43-0.63) and 0.51 (95% CI: 0.44-0.59), respectively, which were significantly lower.^3^ In the matched case-control method, the odds of secondary infection among contacts of AZ and PfBnT vaccinated individuals were also significantly lower, 0.62 (95% CI: 0.48-0.79) and 0.51 (95% CI: 0.42-0.62) respectively.^3^ The baseline serology and PCR of household contacts were not reported (Table 8).

### Vaccine Efficacy or Effectiveness Against Asymptomatic Infection

Ten studies reported vaccine efficacy or effectiveness against asymptomatic COVID-19 infection (Table 9 and Table 10). Three of these involved AZ vaccine,^4–6^ another four examined PfBnT vaccine^8, 11, 12, 38^ and one study each evaluated mRNA-1273^18^ and J and J ^19^ vaccines and one evaluated both mRNA-1273 and BNT162b2.^10^ The methods of assessing efficacy or effectiveness against asymptomatic infection used in some of these studies include RT-PCR nasopharyngeal swabs at time intervals.

**Table 9:** First-dose Vaccine Efficacy or Effectiveness Against Asymptomatic Infection

| Vaccine | Author | Country | Strain targeted by PCR | Baseline Serology | Dosing Schedule | Follow-up days* | Outcomes | Vaccine Efficacy or effectiveness (95%CI)<sup>+</sup> |
| --- | --- | --- | --- | --- | --- | --- | --- | --- |
| **AstraZeneca**<br>(ChAdOx1 nCoV-19) | Voysey et al.<sup>5</sup> (RCT) | UK | Wild type | Negative | LD or SD | >21 | Asymptomatic or unknown | 7.8% (-46.7-42.1) |
|  | Voysey et al.<sup>5</sup> | UK | Wild type | Negative | LD or SD | >21 | Any PCR+ | 46.3% (31.8-57.8) |
|  | Voysey et al.<sup>4</sup> (RCT) | UK/Brazil/S.Afrca | Wild type | Negative | LD or SD | 22-90 | Any PCR+ | 67% (49-78) |
|  | Voysey et al.<sup>4</sup> | UK/Brazil/S.Afrca | Wild type | Negative | SD | 22-30 | Asymptomatic or Unknown | 0.2 (-209-68) |
|  | Voysey et al.<sup>4</sup> | UK/Brazil/S.Afrca | Wild type | Negative | SD | 31-60 | Asymptomatic or Unknown | 17% (-172-75) |
|  | Voysey et al.<sup>4</sup> | UK/Brazil/S.Afrca | Wild type | Negative | SD | 22-90 | Asymptomatic or unknown | 16% (-88-62) |
|  | Pritchard et al.<sup>7</sup> | UK | Wild type and B.1.1.7 | Both | NA | 0-7 | Asymptomatic | OR: 0.45(0.35 to 0.57) |
|  | Pritchard et al | UK | Wild type and B.1.1.7 | Both | NA | 8-20 | Asymptomatic | OR: 0.47(0.37 to 0.6) |
|  | Pritchard et al | UK | Wild type and B.117 | Both | NA | ≥ 29 | Asymptomatic | OR: 0.39(0.3 to 0.51) |
| **Janssen Biotech**<br>(Ad26.COV2.S) | Janssen Biotech<sup>19</sup> (Regulatory submission) | Multiple | Wild type | Negative | NA | 1-29 | Asymptomatic | 20% (-7-40.4) |
|  | Janssen Biotech<sup>19</sup> | Multiple | Wild type | Negative | NA | ≥ 29 | Asymptomatic | 74% (46.8-88.4) |
| **Pfizer, BioNTech**<br>(BNT162b2) | Amit et al.<sup>12</sup> | Israel | Wild type | Unknown | NA | 1-14 | Asymptomatic or unknown | NR<sup>##</sup> |
|  | Amit et al.<sup>12</sup> | Israel | Wild type | Unknown | NA | 15-28 | Asymptomatic or unknown | NR<sup>###</sup> |
|  | Dagan et al.<sup>8</sup> | Israel | Wild type and B.1.1.7 | Unknown | NA | 14-20 | Asymptomatic | 29% (17-39) |
|  | Dagan et al.<sup>8</sup> | Israel | Wild type and B.1.1.7 | Unknown | NA | 21-27 | Asymptomatic | 52% (41-60) |
|  | Hall et al.<sup>11</sup> | UK | Wild type | Unknown | NA | 21 days after 1<sup>st</sup> dose and 7 days after 2<sup>nd</sup> | Asymptomatic or unknown | 97.2%<sup>#</sup> |
|  | Haas et al.<sup>9</sup> | Israel | Wild type and B.1.1.7 | Unknown | NA | 14-21 days | Asymptomatic | 52% (48.9–55.0) |
|  | Pritchard et al.<sup>7</sup> | UK | Wild type and B.117 | Both | NA | 0-7 | Asymptomatic | OR: 0.48(0.39 to 0.6) |
|  | Pritchard et al.<sup>7</sup> | UK | Wild type and B.117 | Both | NA | 8-20 | Asymptomatic | OR:0.54(0.45 to 0.65) |
|  | Pritchard et al.<sup>7</sup> | UK | Wild type and B.1.1.7 | Both | NA | ≥ 29 | Asymptomatic | OR: 0.44(0.36 to 0.55) |
|  | Regev-Yochay et al.<sup>14</sup> | Israel | Wild type | Both | NA | 4-10 | Asymptomatic at first test | 28(-18 to 57) |
|  | Regev-Yochay et al.<sup>14</sup> | Israel | Wild type | Both | NA | 4-10 | Asymptomatic (who never became symptomatic) | 27(-38 to 61) |
|  | McEllistrem et al.<sup>37</sup> | USA | Wild Type | Unknown | NA | 12-15 days | Asymptomatic | NR |
|  | Joness et al.<sup>13</sup> | UK | Wild type and B.1.1.7 | Unknown | NA | <12 and >12 | Asymptomatic | NR |
| **Moderna (mRNA-1273)** | Baden et al.<sup>18</sup> (RCT) | USA | Wild type | Negative | NA | From day 1 | Asymptomatic | 61.4%<sup>#</sup> |
| **mRNA vaccines (BNT162b2 or mRNA-1273)** | Tande et al.<sup>10</sup> | USA |  | Unknown | NA | From day 1, at least one dose | PCR+ in asymptomatic | 56% (40-67) |
|  | Bouton et al.<sup>15</sup> | USA | Wild type | Unknown | NA | From day 1, 1 dose | Asymptomatic | RR calculated: -1% (-57-35) |

\* Time PCR was conducted after first or second dose; LD: Low dose, SD: Standard dose, #Calculated from raw values. ## 2.7 cases per 10000 person-days in vaccine vs 2.4 cases per 10000 person-days in control. ###1.7 cases per 10000 person-days in vaccine vs 2.4 cases per 10000 person-days, Calculated from raw values).<sup>+</sup> Efficacy reported for RCTs and Effectiveness for observational studies. All Pfizer BioNtech's studies except Dagan et al. involved healthcare workers. 0.44 (95% CI: 0.33-0.60). Studies are observational except otherwise stated. , <sup>+</sup> VE =1-RR (or HR) x100%

**Table 10:**
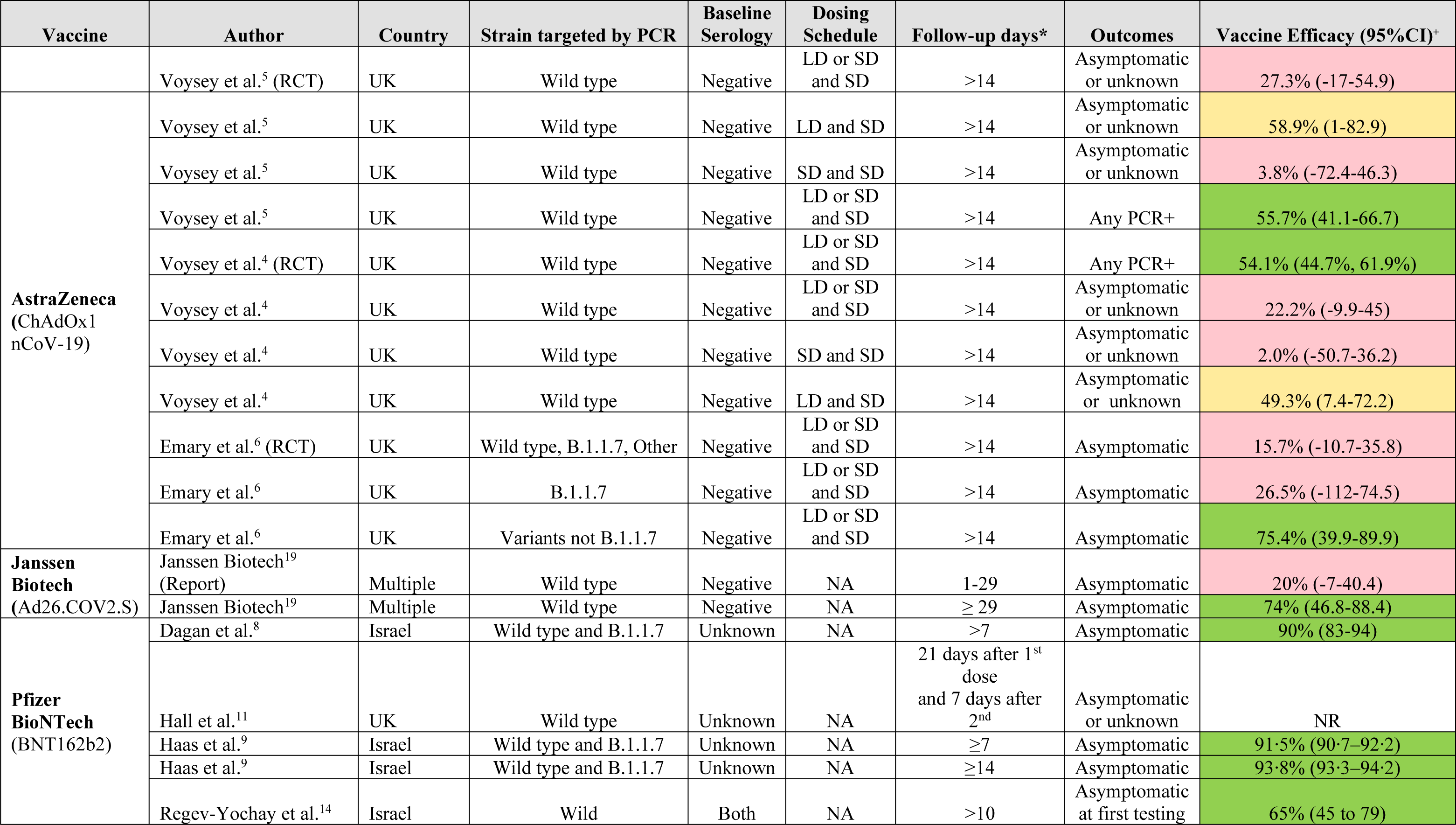
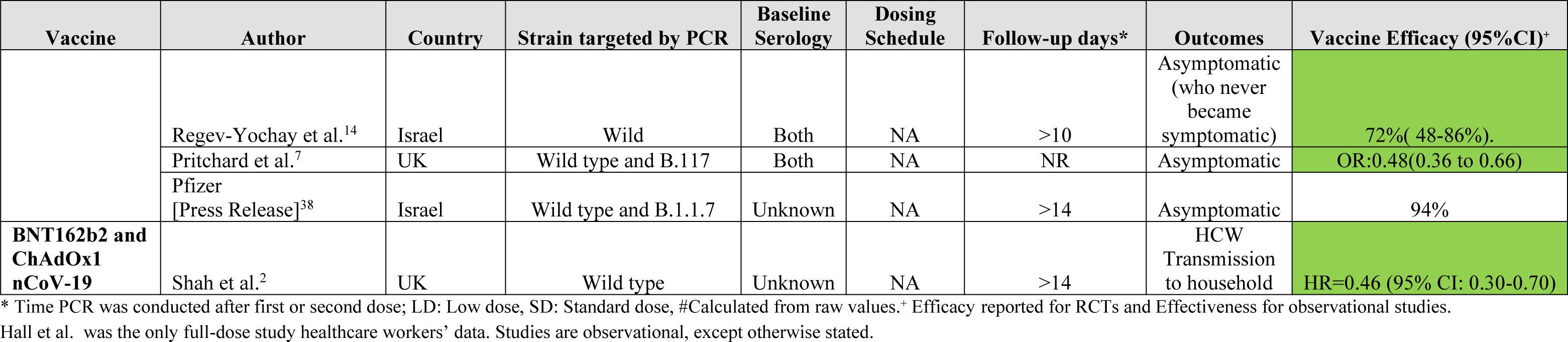
Full-dose Vaccine Efficacy or Effectiveness Against Asymptomatic Infection

#### AstraZeneca Vaccine Efficacy in the General Population

##### First Dose AstraZeneca

Asymptomatic infection data were presented for only the UK component of the AZ vaccine studies. Two AZ vaccine studies reported vaccine efficacy against asymptomatic or unknown infection of 7.8% (-46.7-42.1)^5^ and 16% (-88-62)^4^ respectively, after more than 21 days and 22 to 90 days of the first dose. However, vaccine efficacy among participants with positive results, irrespective of symptoms, were 46.3% (31.8-57.8)^5^ and 67% (49-78)^4^, respectively over the same period (Table 9). These trials implemented weekly self-administered nose and throat swabs for testing on baseline seronegative participants. The PCR status of these participants was not established at baseline.

##### Full Dose AstraZeneca

After 14 days of the second dose, two AZ vaccine studies did not demonstrate efficacy against asymptomatic or unknown infection with the wild type virus 22.2% (-9.9-45) and 27.3% (95% CI: -17-54.9)) respectively.^4, 5^ A third study did not show efficacy against asymptomatic infection with the B.1.1.7 variant (26.5% (95% CI: -112-74.5)), following low or standard dose vaccination.^6^ All three studies involved baseline seronegative participants. The baseline PCR results of the participants were not reported, therefore, persistent carriage after previous infection was not ruled out. In the subgroup of participants with an initial low dose of the vaccine, followed by a standard dose, two studies reported 49.3%(95% CI: 7.4-72.2)^4^ and 58.9%(95% CI: 1-82.9)^5^ respective efficacies against asymptomatic and unknown infection 14 days after the second dose (Table 10).

However, when participants who were PCR positive irrespective of symptoms were considered, two doses of AZ vaccine showed efficacies of 55.7% (41.1-66.7)^5^ and 54.1% (44.7%, 61.9%)^4^ after 14 days of second dose vaccination.

#### AstraZeneca Vaccine Effectiveness in the General Population

##### First Dose AstraZeneca

In a large UK household survey with longitudinal follow-up among seronegative or seropositive individuals, Pritchard et al. reported significant reductions in the odds of asymptomatic infections following AZ vaccine 0-7 days, 8-20 days and 21 or more days after the first dose (ORs: 0.45 (95% CI: 0.35-0.57), 0.47 (95% CI: 0.37-0.6) and 0.39 (95% CI: 0.3

-0.51), respectively).^7^ Nose and throat self-swabs were conducted every week for a month, and subsequently monthly for 12 months from enrolment.^7^

#### Pfizer BioNTech Vaccine Effectiveness in the General Population

##### First Dose Pfizer BioNTech Vaccine

An Israeli observational study by Dagan et al., which did not establish baseline seronegativity, showed that one dose of BNT162b2 significantly reduced asymptomatic infection by 29% (95% CI: 17-39) and 52% (95% CI: 41-60) after 14 to 20 days and 21 to 27 days of follow-up respectively, as assessed by confirmed positive PCR SARS-CoV-2 test without documented symptoms. No routine swabbing was documented for the participants (Table 9).

In a large UK household survey with longitudinal follow-up involving participants with unknown baseline serology status, Pritchard et al. reported significant reductions in the odds of asymptomatic infections following PfBnT vaccine 0-7 days, 8-20 days and 21 or more days after the first dose, ORs: 0.48 (95% CI: 0.39-0.6) and 0.54 (95% CI: 0.45-0.65) respectively, compared with unvaccinated previously PCR negative individuals.^7^ Nose and throat self-swabs were conducted every week for a month, and subsequently monthly for 12 months from enrolment.^7^

##### Full Dose Pfizer BioNTech Vaccine

Dagan et al. also demonstrated 90% effectiveness (95% CI: 83-94) against asymptomatic infection seven days after the second dose.^8^ A press release by the vaccine manufacturer reported two weeks post-second dose effectiveness of 94% against asymptomatic infection in Israel.^38^ The study utilized de-identified aggregate Israel Ministry of Health public health surveillance data. The analysis was conducted when more than 80% of tested specimens in Israel were variant B.1.1.7.^38^ In another Israeli study, which utilized the national public health surveillance data, Haas et al. reported significantly higher vaccine effectiveness seven or more days after full dose PfBnT vaccination, 90.4% (95% CI: 89.1-91.5).^9^ The incidence rate per 100 000 person-days among unvaccinated individuals was 54.6 compared with 3.2 in those vaccinated. Vaccine effectiveness after 14 or more days was 93.8% (95% CI: 93.3-94.2).^9^ Pritchard et al. also found full dose vaccination with PfBnT vaccine to significantly reduce the odds of asymptomatic infection compared with unvaccinated previously PCR negative UK residents, 0.48 (95% CI: 0.36-0.66).^7^

#### mRNA (Pfizer BioNTech and Moderna) Vaccines Effectiveness in the General Population

##### First or second dose of mRNA vaccine

Tande et al. evaluated the effectiveness of at least one dose of either mRNA-1273 or PfBnT vaccine among people who underwent molecular tests prior to a procedure or surgery.^10^ The relative risk for a positive test during asymptomatic pre-procedure screening in vaccinated compared with unvaccinated was significantly lower (0.44 (95% CI: 0.33-0.60)). Ten or more days after the 1st dose, the risk of a positive test was also significantly lower among the vaccinated (0.28 (95% CI: 0.16-0.49; p<.0001)). The risk of test positivity was similarly lower among the vaccinated, after the second dose 0.27 (95% CI: 0.12-0.60).^10^

#### Moderna Vaccine Efficacy in the General Population

##### First Dose Moderna Vaccine

A study of the mRNA-1273 reported that 0.1% of the participants receiving the first dose developed asymptomatic infection, assessed at second dose with nasal swabs, compared with 0.27% of the unvaccinated group, 21 days after the first dose; which is suggestive of 61.4% efficacy against asymptomatic carriage. Participants in this trial were negative for COVID-19 by RT-PCR or antibody testing at baseline.^18^ There was no data on asymptomatic infection after full dose (Table 9).

#### Janssen Vaccine Efficacy in the General Population

##### Full Dose Janssen vaccine

This is a single dose vaccine. The J&J vaccine did not show statistically significant efficacy against asymptomatic infection in the first 29 days of follow-up. However, after 29 days post-vaccination, asymptomatic infection, assessed via surveillance swabs at unspecified intervals among baseline seronegative participants, was significantly lower among vaccinated participants (74%, 95% CI: 46.8-88.4%).^19^ Asymptomatic infection in this trial was assessed by lack of symptoms on the day preceding, the day of, or any time after a positive PCR test. Furthermore, efficacy as demonstrated by seroconversion in previously asymptomatic participants was 74.2% compared with placebo (95% CI: 47.1; 88.6).^19^

#### Effectiveness of Vaccines in Health Care Workers (HCWs)

Hall et al. in a surveillance study of HCWs with documented baseline PCR and antibody in the UK, reported a higher incidence density of asymptomatic or unknown test positivity in unvaccinated HCWs than PfBnT or AZ vaccine recipients. In this biweekly surveillance of vaccine status, symptoms, and nasal or nasal plus oral swabs, the vaccine recipients were followed up for 396,318 person-days compared with 710,587 person days in unvaccinated group, with 35 and 218 asymptomatic or unknown infections reported, translating to 0.88 and 3 infections per 10,000 person-days respectively.^11^The vaccine effectiveness overall was 72% from 21 days after dose one and 86% after two. Protection was noted from day 10 post first dose in these data. The study took place over two months and involved 23,320 HCWs in 104 hospitals, of whom 35% were previously documented SARS-CoV-2 positive. Also, of practical relevance in those testing positive, the unvaccinated group had a higher proportion with “classic” symptoms at 63%: 14% had other symptoms, 5% were asymptomatic and 17% unknown. In the vaccinated group only 40% had classic symptoms, 13% had other symptoms, 13% were asymptomatic, and 31% had unknown – therefore asymptomatic or non-classic symptoms comprised 26% of the post vaccine test positive. In a second observational HCW study in Israel by Amit et al.,^12^ asymptomatic test positive data was limited, and reasons for tests done without documented symptoms was not reported. The baseline RT-PCR status was not assessed; therefore, prolonged RT-PCR positivity after prior infection was not ruled out. Estimated asymptomatic infection prevalence in the vaccinated group was 1.7 per 10,000- person years between 15 and 28 days after vaccination, compared with 2.4 per 10,000 person-years in the unvaccinated group (VE: 29%).^12^ The overall infection rate in the whole cohort for the 2-month study was 1.9%. Matheson et al. in an asymptomatic weekly screening study among HCWs who were vaccinated with one dose of PfBnT vaccine compared with unvaccinated HCWs, showed that 0.8% of tests from unvaccinated HCWs were positive compared with 0.37% and 0.2% from vaccinated ones at <12 days and >12 days post-vaccination respectively (p=0.023 and p=0.004, respectively).^13^

In another large Israeli cohort study of HCWs who were either seropositive or negative at baseline, Regev-Yochay et al.^14^ reported a prevalence of PCR positive asymptomatic (at first testing) infection following exposure to COVID-19 of 5.2% among unvaccinated HCW compared with 1.8% among fully PfBNT vaccinated HCW (VE:65%, 95%CI: 45-79%). The prevalence of infection among true asymptomatic (who never became symptomatic) was 3.3% among unvaccinated and 0.9% among vaccinated individuals. The vaccine effectiveness was 72% (95% CI: 48-86%).^14^ Furthermore, the vaccine was 70% effective (95% CI: 43-84%) in preventing all presumably infectious cases (with Ct values <30) and 83% effective (95% CI: 51-94%) against presumably infectious asymptomatic cases.^14^ Bouton et al. however did not find any significant difference in asymptomatic cases (assessed following workplace exposure, out of state travel or upon request) at any time from the first dose of vaccination with either Moderna or PfBnT vaccines compared with unvaccinated HCWs at a tertiary health centre.^15^

### Risk of Vaccinated HCW Developing COVID-19

Table 11 to Table 13 utilized estimates from the current HCW specific literature on likelihood of asymptomatic, symptomatic or any infection over a range of attack rates (to encompass a range of exposure risk) to describe the risk of vaccinated HCW developing COVID-19 after a potential exposure to support decision makers in policy design. The risk reduction values used are from an evolving body of literature and a range of study designs, with reduction of risk noted by weeks from first dose as extracted from the studies. Vaccine related risk reduction ranges are based on available HCW study data, from the following time points: 3 weeks post dose 1 mRNA; 3 weeks to 12 weeks post dose 1 nonreplicating adenovirus. Vaccination protection following first dose was selected both based on available data and because this is relevant as the dosing schedules are currently based on an extended interval for many vaccinees. These data should be considered within the context of current uncertainties including long-term follow up data beyond three months after vaccination.

**Table 11:** Risk of First Dose Vaccinated HCW Developing Asymptomatic COVID-19 (Approximately 20% of All Cases) After a Potential Exposure (Data Estimates from Wild Type SARS-CoV-2)^1^

| **Attack Rate based on Exposure Risk<sup>2</sup>** | **Risk of Developing COVID-19 based on Proportion of Type of COVID-19 Infection x Exposure Risk** | **Vaccination Type** | **Vaccine Risk Reduction (assuming risk reduction [protection] remains the same regardless of exposure risk)<sup>3</sup>** | **Risk of developing COVID-19 following vaccination regimen** |
| --- | --- | --- | --- | --- |
| *A* | *B*<br>(*A* x 20%) |  | *C* | *D*<br>(*B* x *C*) |
| Low risk<br>0.5% | 0.1% | mRNA vaccine | 0.39 - 0.03<br>(61%-97% 3 weeks after first dose) | **1/2,564 – 1/33,333** |
|  |  | Nonreplicating adenovirus vaccine | 0.25<br>(75% 12 weeks after first dose) | **1/4,000** |
| Medium risk<br>1% | 0.2% | mRNA vaccine | 0.39 - 0.03<br>(61%-97% 3 weeks after first dose) | **1/1,282 - 1/16,666** |
|  |  | Nonreplicating adenovirus vaccine | 0.25<br>(75% 12 weeks after first dose) | **1/2,000** |
| High risk<br>5% | 1% | mRNA vaccine | 0.39 - 0.03<br>(61%-97% 3 weeks after first dose) | **1/256 - 1/3,333** |
|  |  | Nonreplicating adenovirus vaccine | 0.25<br>(75% 12 weeks after first dose) | **1/400** |
| Superspreader event<br>10% | 2% | mRNA vaccine | 0.39 - 0.03<br>(61%-97% 3 weeks after first dose) | **1/128 - 1/1,666** |
|  |  | Nonreplicating adenovirus vaccine | 0.25<br>(75% 12 weeks after first dose) | **1/200** |

2 Estimates for attack rate derived from study data and stratified to estimate range from low risk to superspreader event risk. from Abbas et al. which identified HCW risk to range from ~2-8%. Abbas, M., Robalo Nunes, T., Martischang, R. et al. Nosocomial transmission and outbreaks of coronavirus disease 2019: the need to protect both patients and healthcare workers. Antimicrob Resist Infect Control 10, 7 (2021). https://doi.org/10.1186/s13756-020-00875-7
3 Study data applied to all infections and asymptomatic infections: nonreplicating adenovirus vaccines - AZ: 16% after 3 weeks, (25-60% after 12 weeks, NS); Janssen: 75% after 12 weeks (PCR and/or serology without previous symptoms). mRNA vaccines - Moderna: 61% from 3 weeks after dose 1; Pfizer BioNTech (Hall et al): 97% 21 days after dose 1; Pfizer BioNTech (Daagen et al): 29% effectiveness 15 days after dose 1 in HCW

**Table 12:** Risk of Vaccinated HCW Developing Symptomatic COVID-19 (80% of All Cases) After a Potential Exposure (Data Estimates from Wild Type SARS-CoV-2) Compared with No Vaccination^1^

| Attack Rate based on Exposure Risk<sup>2</sup><br>*A* | Risk of Developing COVID-19 based on Proportion of Type of COVID-19 Infection x Exposure Risk<br>*B*<br>(*A* x 80%) | Vaccination Type<br>Nonreplicating adenovirus vaccine or mRNA vaccine | Vaccine Risk Reduction, time after first dose by vaccine type (assuming risk reduction [protection] remains the same regardless of exposure risk)<sup>3</sup><br>*C* | Risk of vaccinated person developing COVID-19 by regimen<br>*D*<br>(*B* x *C*) |
| --- | --- | --- | --- | --- |
| Low risk<br>0.5% | 0.4% | Nonreplicating adenovirus vaccine effectiveness range across studies (67% 3w post dose 1 to -75% 12 w post dose 1) | 0.22 to 0.66 | **1/757**<br>**to**<br>**1/1,136** |
|  |  | mRNA vaccine effectiveness range (70% 3 w post dose 1 to 95% 2 wks post dose 1) | 0.3 to 0.05 | **1/833**<br>**to**<br>**1/5,000** |
| Medium risk<br>1% | 0.8% | Nonreplicating adenovirus vaccine effectiveness range | 0.33<br>(67% effectiveness 3 weeks post first dose) | **1/378** |
|  |  |  | 0.22<br>(75% maximum reduction from 12 weeks onwards) | **1/568** |
|  |  | mRNA vaccine effectiveness range | 0.3<br>(70% reduction 3 weeks after first dose) | **1/416** |
|  |  |  | 0.05<br>(95% reduction 2 weeks after first dose) | **1/2,500** |
| High risk<br>5% | 4% | Nonreplicating adenovirus vaccine effectiveness range | 0.33<br>(67% effectiveness 3 weeks post first dose) | **1/75** |
|  |  |  | 0.22<br>(75% maximum reduction from 12 weeks onwards) | **1/113** |
|  |  | mRNA vaccine effectiveness range | 0.3<br>(70% reduction 3 weeks after first dose) | **1/83** |
|  |  |  | 0.05<br>(95% reduction 2 weeks after first dose) | **1/500** |
| Superspreader event<br>10% | 8% | Nonreplicating adenovirus<br>vaccine effectiveness range | 0.33<br>(67% effectiveness 3 weeks post first dose) | **1/37** |
|  |  |  | 0.22<br>(75% maximum reduction from 12 weeks onwards) | **1/56** |
|  |  | mRNA vaccine effectiveness<br>range | 0.3<br>(70% reduction 3 weeks after first dose) | **1/41** |
|  |  |  | 0.05<br>(95% reduction 2 weeks after first dose) | **1/250** |

<sup>1</sup> Estimate of type of COVID-19 cases (symptomatic ~80%) is a conservative estimate derived from a meta-analysis on the proportion of asymptomatic case and potential range for community transmission. Byambasuren O, Cardona M, Bell K, Clark J, McLaws M-L, Glasziou P. Estimating the extent of asymptomatic COVID-19 and its potential for community transmission: systematic review and meta-analysis. Official Journal of the Association of Medical Microbiology and Infectious Disease Canada. 2020;5(4):223-234. https://jammi.utpjournals.press/doi/pdf/10.3138/jammi-2020-0030
<sup>2</sup> Estimates for attack rate derived from study data and stratified to estimate range from low risk to superspreader event risk. from Abbas et al. which identified HCW risk to range from ~2-8%. Abbas, M., Robalo Nunes, T., Martischang, R. et al. Nosocomial transmission and outbreaks of coronavirus disease 2019: the need to protect both patients and healthcare workers. Antimicrob Resist Infect Control 10, 7 (2021). https://doi.org/10.1186/s13756-020-00875-7
<sup>3</sup> Study data applied to all infections and asymptomatic infections: nonreplicating adenovirus vaccines - AZ: 16% after 3 weeks, (25-60% after 12 weeks, NS); Janssen: 75% after 12 weeks (PCR and/or serology without previous symptoms). mRNA vaccines - Moderna: 61% from 3 weeks after dose 1; Pfizer BioNTech (Hall et al): 97% 21 days after dose 1; Pfizer BioNTech (Dagen et al): 29% effectiveness 15 days after dose 1 in HCW

**Table 13:**
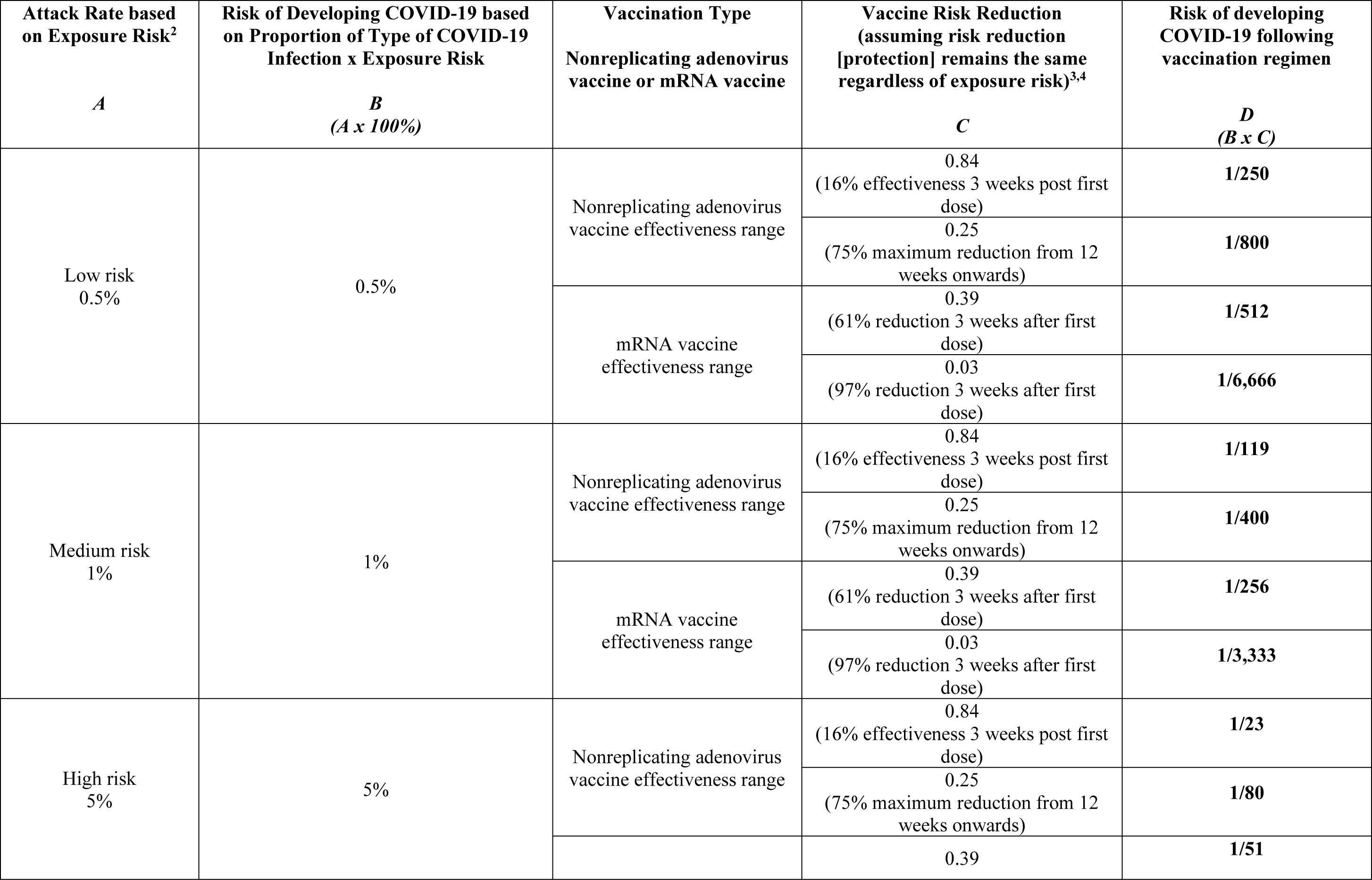
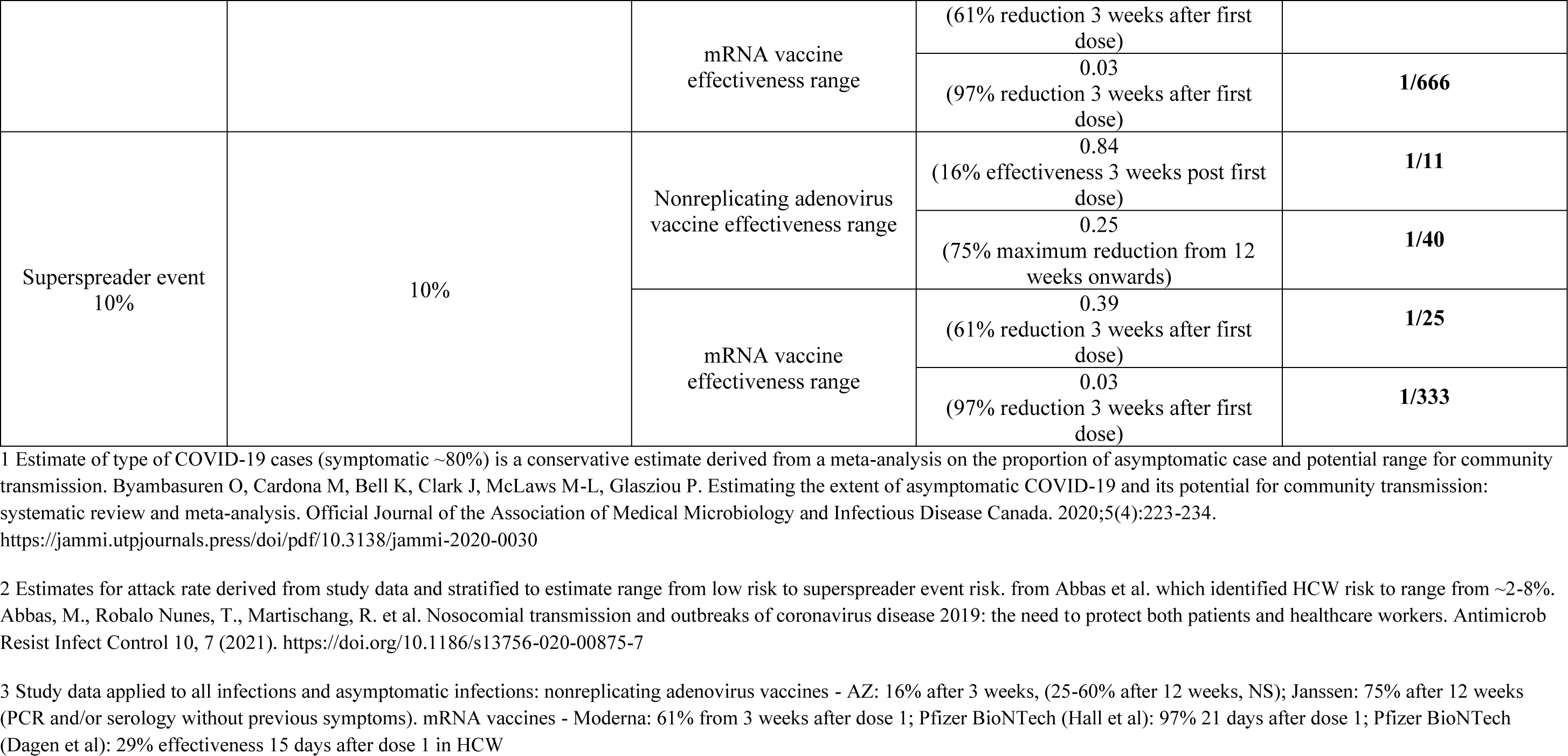
Estimated Risk of Vaccinated HCWs Developing Symptomatic or Asymptomatic COVID-19 After Varied Risk Exposures (Current Data Estimates from Wild Type SARS-CoV-2 and Are Evolving - Will Require Frequent Updates) Compared with No Vaccination^1^

As an example, the risk of a HCW developing an asymptomatic COVID-19 infection (estimated to be 20% of all infections) following a low-risk exposure (0.5%) after receiving first dose of vaccine would be between 1/757 (3 weeks post vaccination) and 1/1,136 (12 weeks post vaccination) for adenovirus vector vaccines and 1/833 to 1/5,000 3 weeks post vaccination for mRNA vaccines.

### Cycle Threshold (Ct) Values

Eight studies reported on Ct, an inverse proxy for viral load. Five of these are new to the updated version of this report.

Results from Phase 2/3 vaccine efficacy studies of AZ vaccine compared with a comparator meningococcal vaccine in the United Kingdom, showed that the Ct values in infected vaccinated participants were statistically significantly higher than the comparator (p<0.0001), after 14 days of the second dose in baseline seronegative efficacy cohorts.^6^ Furthermore, the vaccine recipients were PCR-positive for a significantly shorter period of time (p<0.0001). The Ct values in asymptomatic cases were also significantly higher among vaccine recipients than control (p=0.0040); however, this difference was not significant for primary symptomatic cases (p=0.1534). Vaccine recipients infected with the B.1.1.7 variant also showed significantly higher Ct values than control (p=0.0113).^6^

A longitudinal UK household survey by Pritchard el al. found statistically significant increase in the median Ct values of PfBnT or AZ single or full dose vaccinated individuals compared with unvaccinated individuals at any time point before or after 21 days post-vaccination (p<0.001).^7^ Similarly, in another UK study by Shrotri et al., the mean Ct value of unvaccinated individuals within 27 days of vaccination was 26.6 (95% CI: 26-27.1) compared with 26·6 (95% CI: 25.19-26.62) with one dose of PfBnT or AZ, which was not significantly different (p=0.158).^16^ However, after 28 days, there was a statistically significant decrease in the mean Ct between vaccinated and unvaccinated persons (mean Ct 26.6 (95% CI: 26-27.1) vs 31·3 (95% CI: 29.6-32.9), p<0·001).^16^ Monthly routine PCR testing was conducted in these patients; however, the baseline serology was not reported.^16^ In a longitudinal cohort study of HCWs who were offered voluntary nasal and oropharyngeal swab PCR testing every two weeks as well as serological testing, Lumley et al., found vaccination with either PfBnT or AZ to non-significantly increase Ct value by a mean of 2.7.^17^

A retrospective study of PfBnT mRNA vaccine recipients compared with demographically matched control group of unvaccinated individuals in Israel, found no significant differences in the Ct values for any of the 3 genes (RdRp, N and E) measured less than 12 days after the first dose in infected persons. However, between 12 and 28 days after the first dose, the Ct values for the 3 genes were significantly higher among infected vaccinated persons than controls (p<10^-8^).^36^ In another UK study of one dose of BNT162b2 vaccine, the median Ct values of infected HCWs were reported to have shown a non-significant trend towards increase between unvaccinated (Median=20.3) and vaccinated HCWs after 12 days post-vaccination (Median=30.3), suggesting that samples from infected vaccinated individuals had lower viral loads.^13^ A study by McEllistrem et al. among Community Living Centre residents reported five cases of asymptomatic infections (determined by surveillance nasal swabs every 2-5 days) among baseline PCR negative PfBnT vaccinated and unvaccinated residents. The median Ct values among unvaccinated residents (12.8, IQR: 12.4-14.9) were significantly lower (p=0.009) than vaccinated residents (19.4, IQR: 18.9-25.5).^37^ Furthermore, viral load was -2.4 mean log10 lower among the vaccinated cohort (p=0.004).^37^ In another large cohort study of HCWs at a large medical centre in Israel by Regev-Yochay et al, the mean Ct values among

PfBNT fully vaccinated HCWs (27.3±2.2) was significantly higher (mean difference 5.09, 95% CI: 2.8-7.4, p<0.001) than unvaccinated HCWs (22.2±1.0).^14^

### Animal Studies

All the animal studies estimated viral load by using viral genomic RNA (gRNA) and subgenomic RNA (sgRNA), from nasal or bronchoalveolar lavage (BAL) samples. The vaccines have either been approved or have progressed to human trials.

#### Adenovirus Based Vaccines

van Doremalen et al. found that the AZ vaccine significantly reduced viral load in the bronchoalveolar lavage fluid of vaccinated rhesus macaques on the third, fifth and seventh day post viral challenge, compared with control animals.^20^ In the BAL fluid obtained from control animals, viral genomic and subgenomic RNA were detected on all days; while two of the vaccinated animals had detectable viral gRNA three days after challenge. Viral gRNA was detected in nose swabs from all animals and no difference was found on any day between vaccinated and control animals. Mercado et al. reported that all or almost all macaques vaccinated with Ad26-S.PP, an adenovirus serotype 26 (Ad26) vector-based vaccines expressing tPA leader sequence by J&J, had no detectable virus in BAL or nasal samples respectively, compared with sham controls, which showed high titres of viral sgRNA (p<0.0001 and p<0.0001, respectively).^41^

#### DNA Vaccines

Patel et al. found that peak viral sgmRNA and RNA loads in the BAL were significantly lower in INO-4800 vaccinated macaques (two doses) at day 7 post-challenge (peak: p=0.048, day 7: p=0.024).^42^ However, viral sgmRNA was still detectable in the nasal swabs of both the control and INO-4800 vaccinated animals. Viral mRNA levels trended downwards in INO-4800 vaccinated animals.^42^

#### mRNA Vaccines

The mRNA-1273 vaccine showed significantly lower RNA and subgenomic RNA, in BAL and nasal swabs, in the 100-μg dose group compared with the control group.^21^ Only one of the eight vaccinated had detectable subgenomic RNA in BAL fluid two days after challenge, while all the control animals had detectable RNA. Similarly, none of the eight animals administered the 100-μg dose had detectable subgenomic RNA detected in nasal swab compared with six of eight in the control. Ji et al. found no viral RNA in the lung tissues of mice 42 days after a second dose of BNT162b2-vaccination, compared with a mean of 10^6^ copies/mL in control animals (p<0.001).^40^ A study by Vogel et al. showed BNT162b2-immunized, SARS-CoV-2 challenged macaques cleared all viral RNA by day 3 after challenge; while viral RNA was detected in bronchoalveolar lavage fluid from seven of nine control macaques. One day post challenge, 4 of 9 control animals had detectable viral RNA. In subsequent nasal swabs, viral RNA was detected from some of the control macaques at each sampling time point (5 of 9 on day 3; 4 of 9 on day 6; and 2 of 9 on days 7–23), but none of the BNT162b2-immunized macaques at any sampling time point.^44^

#### Recombinant Nanoparticle Vaccine

In the Novavax’s NVX-CoV2373 vaccine study, macaques administered placebo had elevated viral load two and four days post viral challenge, while all but one of the vaccinated animals had no detectable sgRNA in their BAL fluid.^22^ Similarly, half of the placebo group had elevated viral RNA in their nasal swabs, while none was detectable in vaccinated animals.^22^ Gorman et al. in a macaque study of the 5μg or 25μg doses of NVX-CoV2373 vaccine, found the highest levels of viral sgRNA in placebo animals across the upper and lower-respiratory tract samples, compared with vaccinated animals. By day 8, sgRNA was undetectable in the BAL of animals vaccinated with either doses.^39^ Similarly, Tian et al. showed that placebo-treated mice had an average of 10^4^ SARS-CoV-2 pfu/lung, while those vaccinated with NVX-CoV2373 without Matrix-M had 10^3^ pfu/lung and those with Matrix-M had limited to no detectable virus load.^43^

#### Inactivated Virus-based Vaccines

After two doses of BBV152, an inactivated virus-based COVID-19 vaccine, Yadav et al. detected gRNA and sgRNA in the BAL, nasal and throat swab specimens of all or almost all the macaques in the placebo group by the 7^th^ day post infection, while vaccinated animals had no detectable virus by day 7.^46^ Full dose Sinopharm’s BBIBP-CorV vaccine was reported by Wang et al. to clear COVID-19 virus from the lungs and throats of vaccinated macaques, which were significantly different from the results in the placebo group.^45^ Another study showed that PiCoVacc, a vaccine developed by Sinovac Biotech (China), significantly reduced viral load in the pharynx and lungs of all the vaccinated animals compared with control animals, on the third and seventh day post-viral challenge.^49^

#### Others

Following vaccination with a single dose of recombinant vesicular stomatitis virus-based vaccine (rVSV-ΔG-spike vaccine), Yahalom-Ronen et al. found infectious SARS-CoV-2 in the lungs of infected unvaccinated golden Syrian hamsters, with an average viral titre of 1.3 × 10^5^ pfu/lung, compared with viral titres of 10^4^ -10^8^ pfu in the lungs of animals vaccinated with varying doses of the vaccine, which were below the limit of detection.^47^

## Discussion

In this update, 16 additional studies, including 9 human and 7 animal studies, were included. Therefore, this review has a total of 33 included studies, twenty-one of which were in humans and 12 were preclinical animal studies. Two new studies from Scotland and England evaluated household transmission following vaccination and found PfBnT and AZ to significantly reduce the risk of household transmission.^2, 3^ The majority of the vaccines included in this review demonstrated efficacy and effectiveness against asymptomatic wild-type COVID-19 infections.

The AZ and PfBnT vaccines were found to be significantly associated with higher Ct values than their respective comparators, suggesting that these vaccines may potentially reduce viral load and consequently lower the risk of transmission. It is however noteworthy that the relationship between viral load, viral shedding, infectivity and the duration of infectivity are not well understood. Ct values are also subject to error.^50^

Preclinical primate studies showed that vaccinated animals receiving the Moderna mRNA (8 weeks prior),^21^ PfBnT, or Novavax protein vaccine (35 days prior),^22^ were less likely to have the virus recovered from nasal or lower respiratory samples than unvaccinated animals. The AZ vaccine was more protective against lower respiratory replication, while showing no difference in nasal virus replication.^20^

Studies suggesting the plausibility of vaccine-induced reduction of transmission, including monoclonal antibody therapeutics trials and epidemiologic evidence of transmission from individuals who were persistently RT-PCR positive after natural infection, with evidence of an immunologic response, indicated that viral load can be reduced by circulating antibodies, and that a lower viral load or higher Ct on RT-PCR was associated with a reduced risk of transmission.^25, 26^ However, RT-PCR positivity in the presence of neutralizing antibody and or correlates of cell mediated immunity should not be considered to necessarily represent transmissible infection.

There were significant limitations to many of the included studies. The primary endpoint of the vaccine randomized controlled trials were detection of test positive symptomatic COVID-19, however some studies presented data, which suggest a reduction in the likelihood of testing positive for SARS-CoV-2 RT-PCR in the absence of documented symptoms after vaccination. Furthermore, it was not possible to directly compare findings across studies owing to variations in the assessment of symptom status, and the testing used and timing of these assessments. Also, the possibility of persistent PCR positivity after COVID-19 infection^51^ could not be excluded in some of the studies without baseline PCR assessment. Few studies included surveillance nasal swabs for PCR positivity. Most of the current data were around viral detection, rather than evidence of cultivatable virus. Therefore, there was limited data to evaluate the efficacy or effectiveness of COVID-19 vaccines in decreasing viral loads. In addition, there are only a limited number of epidemiologic data addressing evidence of forward transmission after vaccination.

Based on the current evidence, we suggest the following:

1. all vaccinees should self-isolate and seek testing after the development of COVID-19 compatible symptoms
2. Following exposure, the risk of contracting COVID-19 and subsequent forward transmission from asymptomatic or pauci symptomatic viral carriage should be considered in light of whether the exposed individual was vaccinated the time elapsed since immunization and the consequent expected degree of protection on, a case-by-case basis for those in vulnerable setting. When possible, a case-by-case consideration for whether exposed persons are immunized, is necessary. Low-moderate risk exposures could potentially be managed with careful use of personal protective equipment (PPE), and self-monitoring.
3. If a vaccinated HCW is assessed as having a significant exposure before the period of expected robust immunity, high risk exposures may be managed as for unvaccinated persons.
4. All vaccinated persons should continue to use recommended PPE when in close contact with unvaccinated persons.
5. Population and public health data being collected on positive COVID-19 tests occurring after vaccination should be combined with laboratory data on Ct values, identification of variant strain infections, and epidemiologic contact tracing data to prospectively monitor for evidence of forward transmission of infection from vaccinated persons.

## Conclusion

Two months since the publication of the previous version of this report, 16 additional relevant studies have been published. Two of these are large household surveillance studies from the UK suggesting single or full dose of AZ and PfBnT vaccines may prevent household transmission of COVID-19 after 14 days of vaccination. More studies have found the vaccines to significantly reduce the risk of asymptomatic infection and seven of eight studies found significantly increased cycle threshold, suggestive of lower viral load, in AZ or PfBnT vaccinated individuals compared with unvaccinated. Some studies, such as the AZ vaccine RCTs, included data on cross sectional prevalence of positive SARS-CoV-2 RT-PCR from routine swabbing, which suggested efficacy against asymptomatic infection, although this was not routinely assessed in a comparable way across studies. Evidence regarding the Ct values for AZ vaccine and the PfBnT vaccine suggest their potential to reduce viral load and possibly transmission. Further research is needed to evaluate post-vaccination infectivity and transmission of both the wild type COVID-19 virus and the variants of concern from other jurisdictions.

## Data Availability

All datasets supporting the conclusions of this article are included within the article.

## Appendix 1: Search Strategy

Ovid Multifile Database: EBM Reviews - Cochrane Central Register of Controlled Trials <MARCH 2021>, Embase <1974 to 2021 May 03>, Ovid MEDLINE(R) and Epub Ahead of Print, In-Process, In-Data-Review & Other Non-Indexed Citations and Daily <1946 to May 03, 2021>

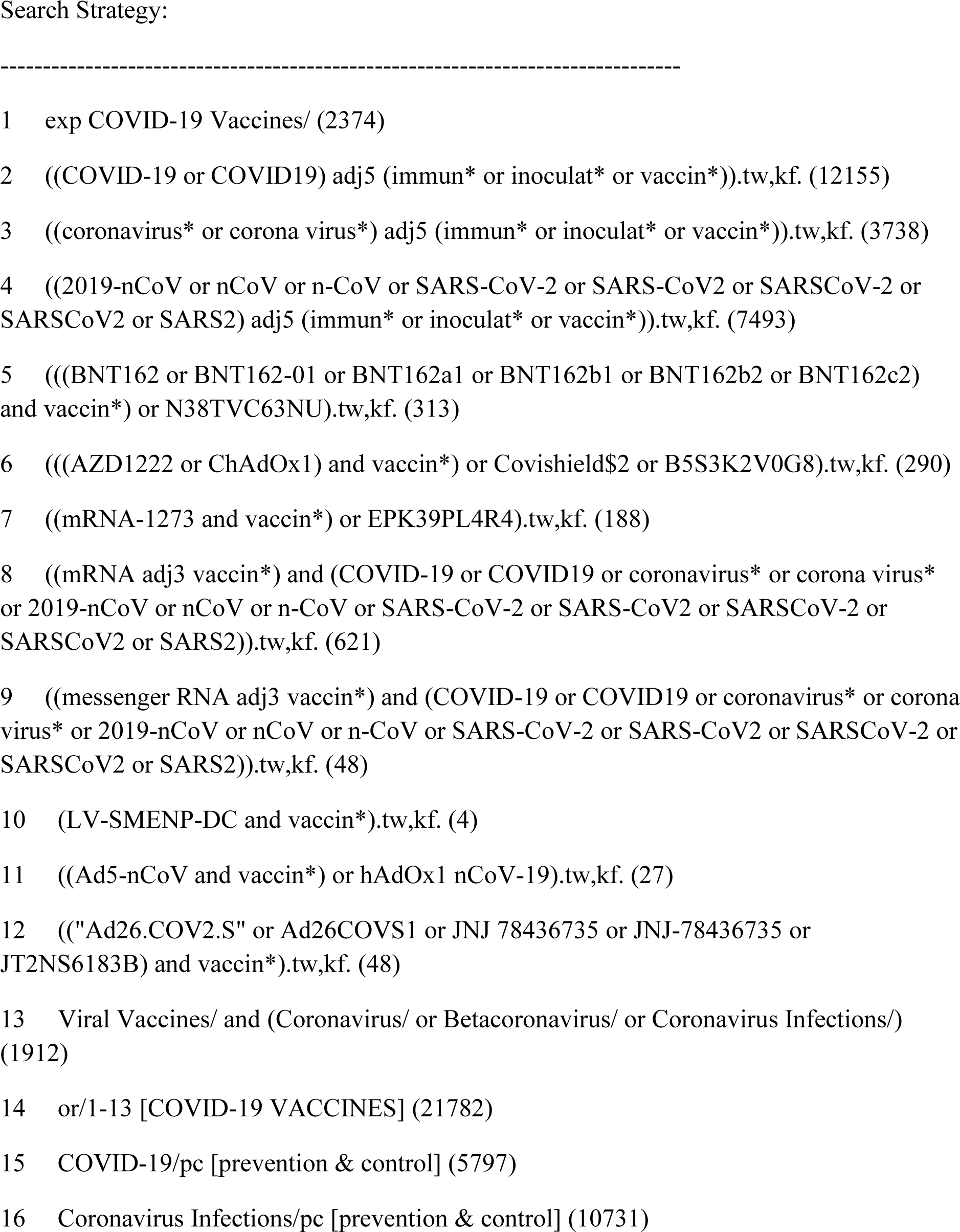
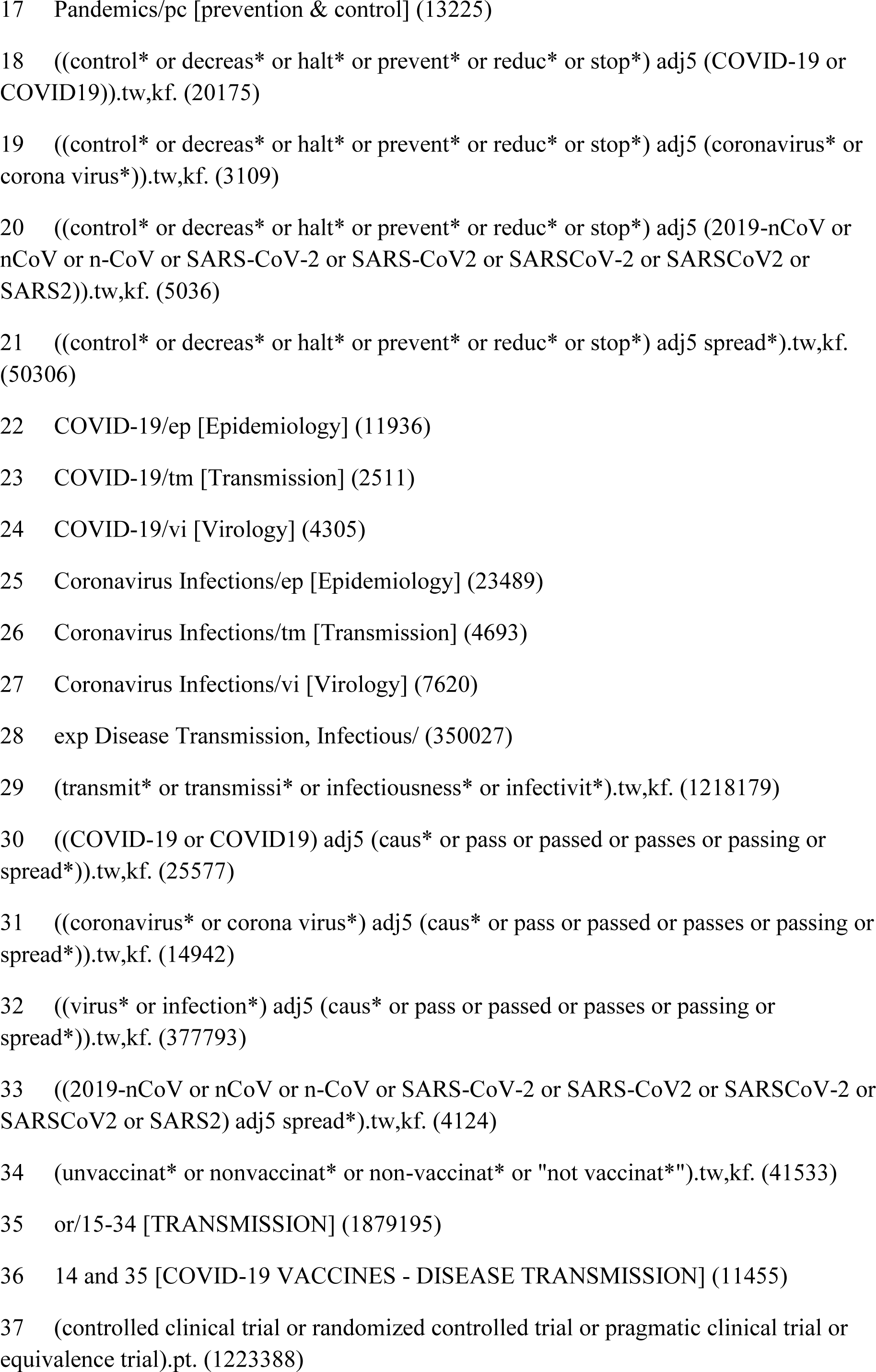
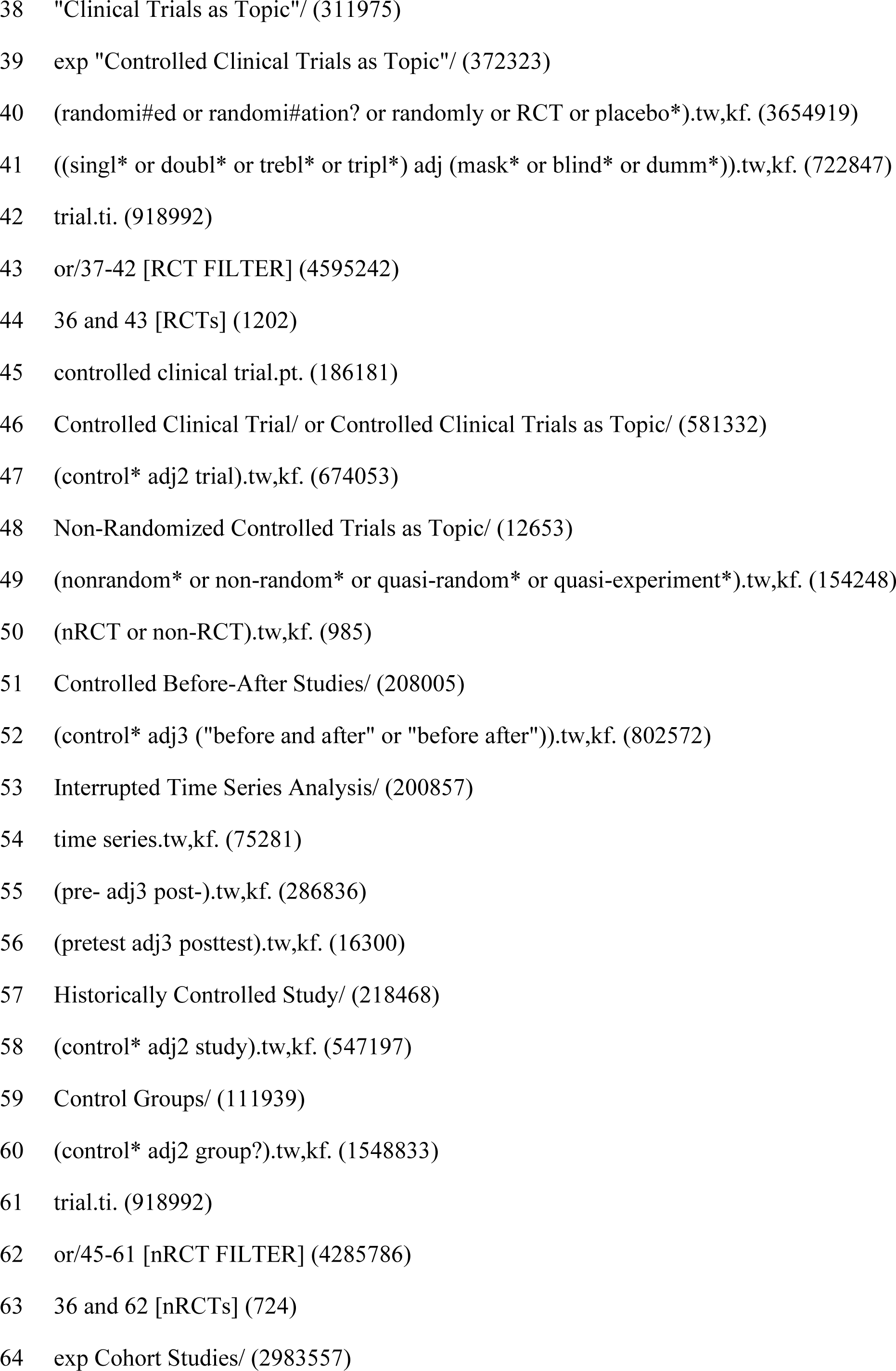
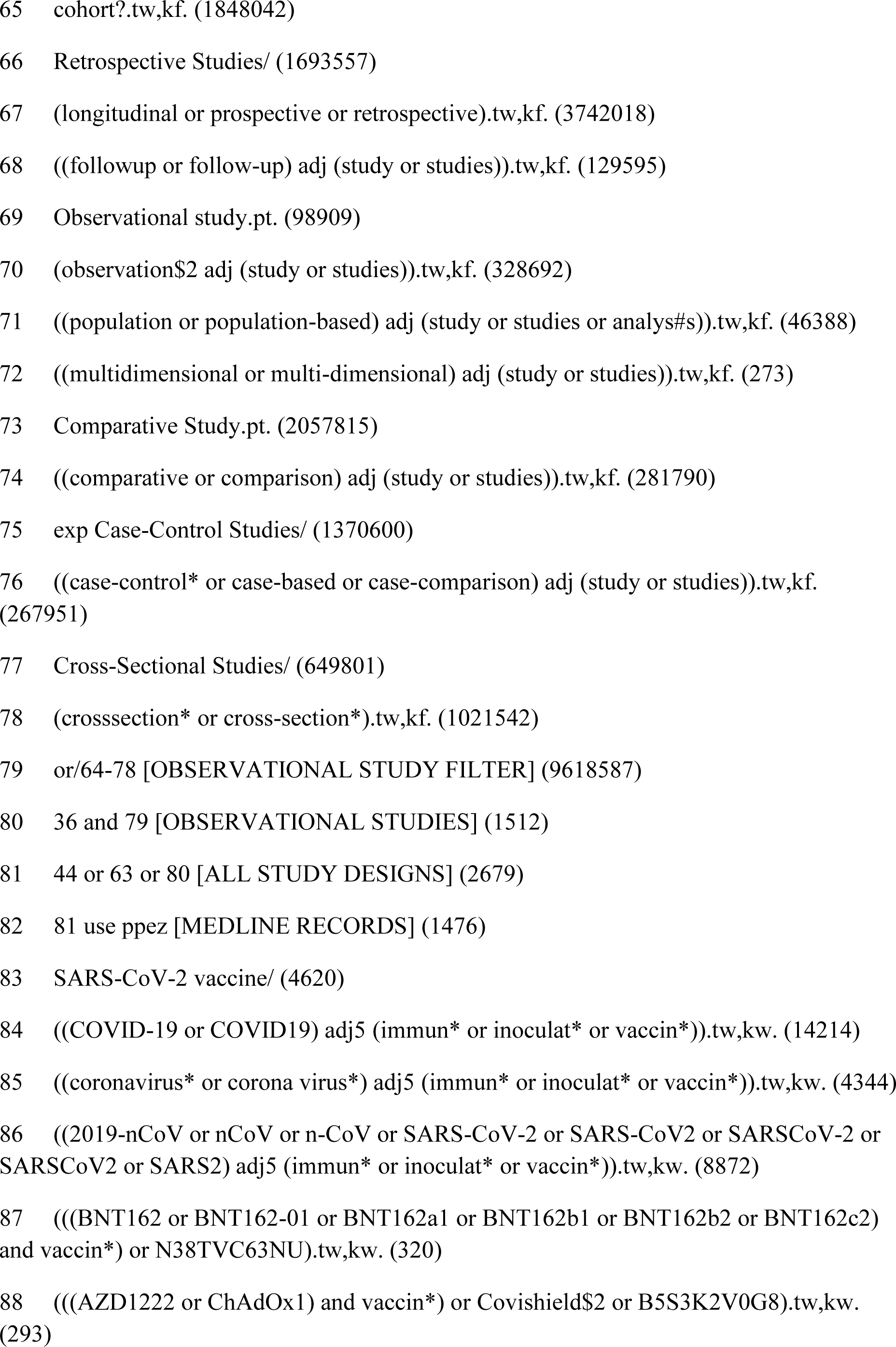
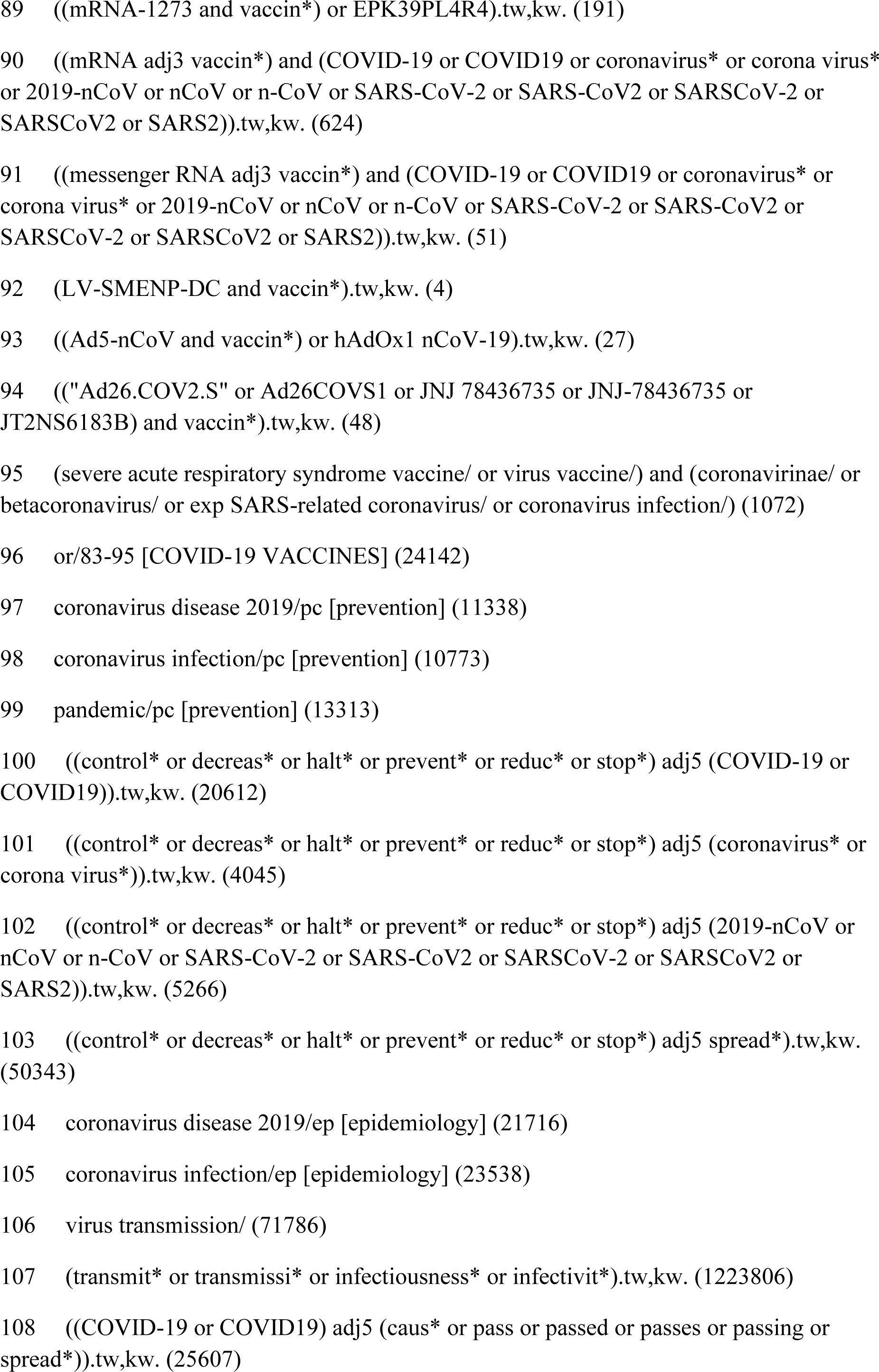
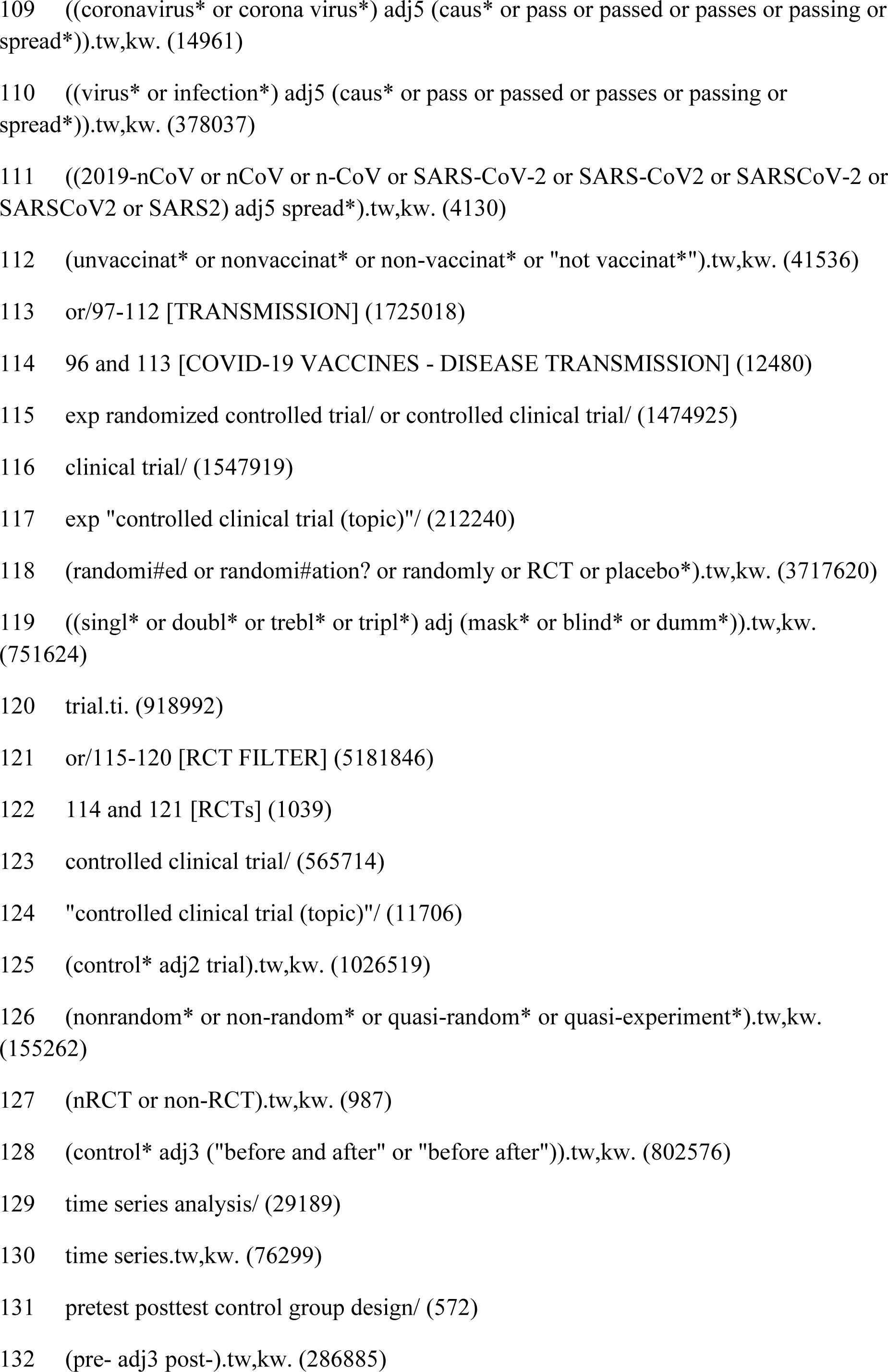
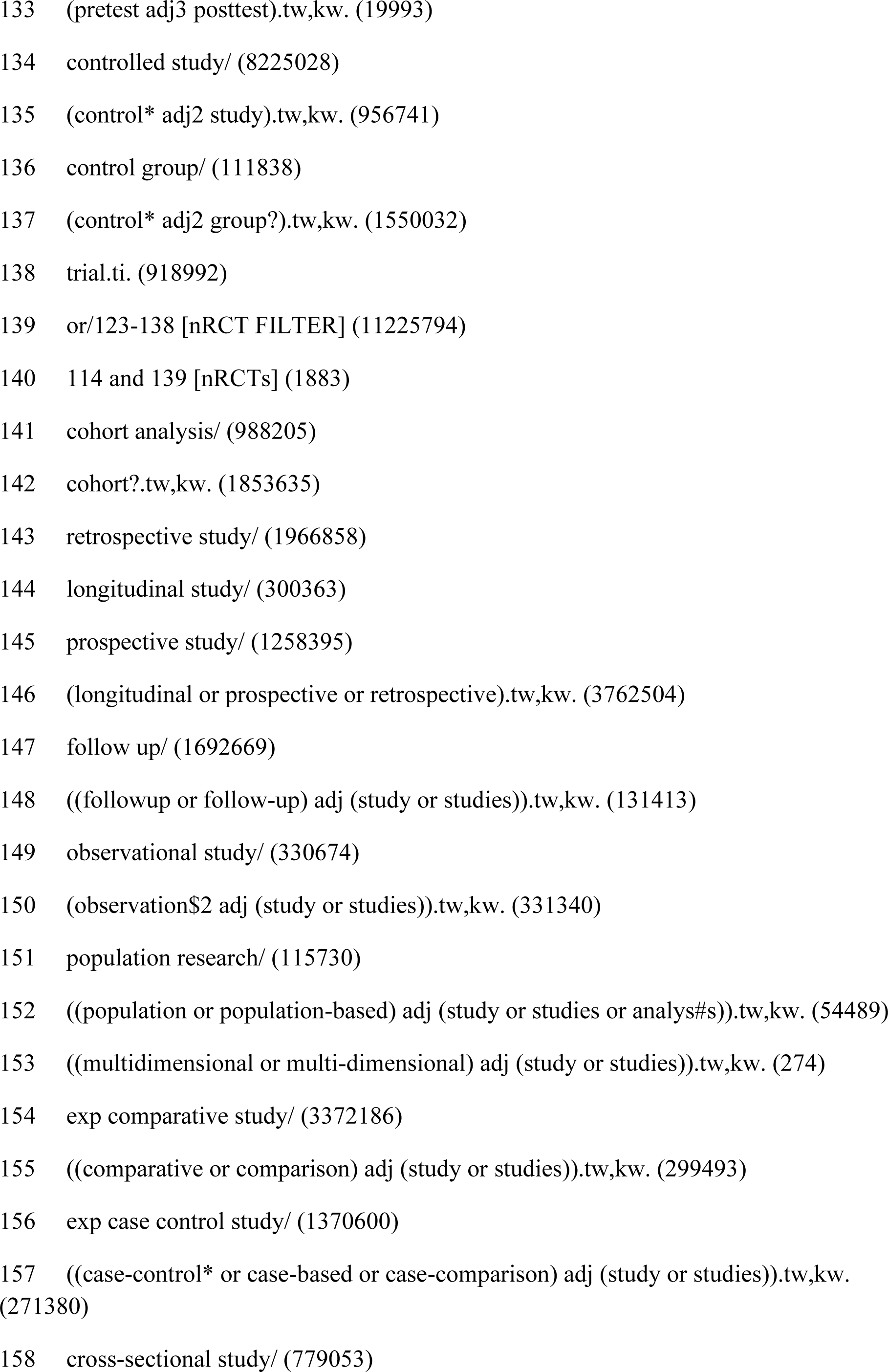
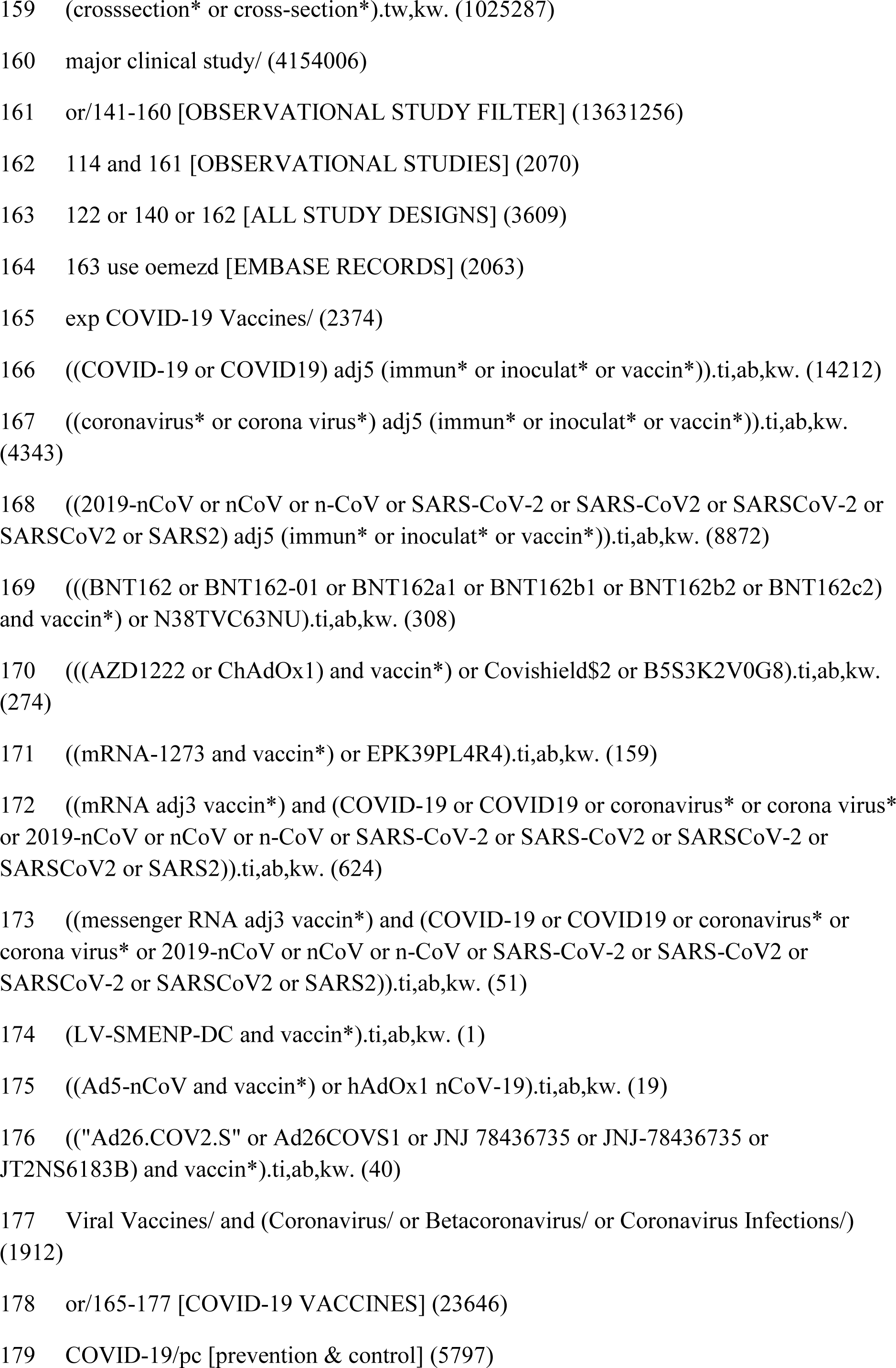
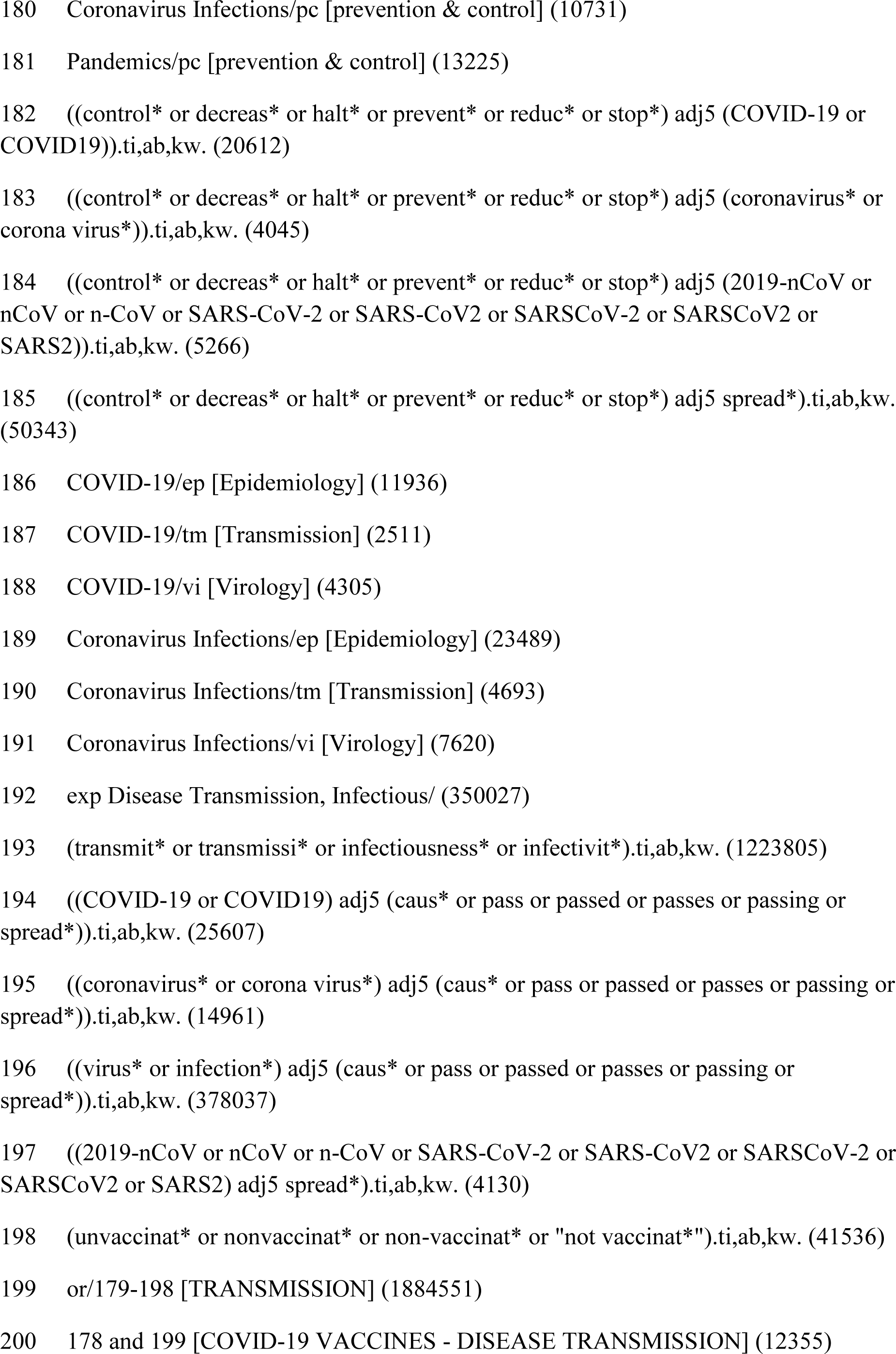
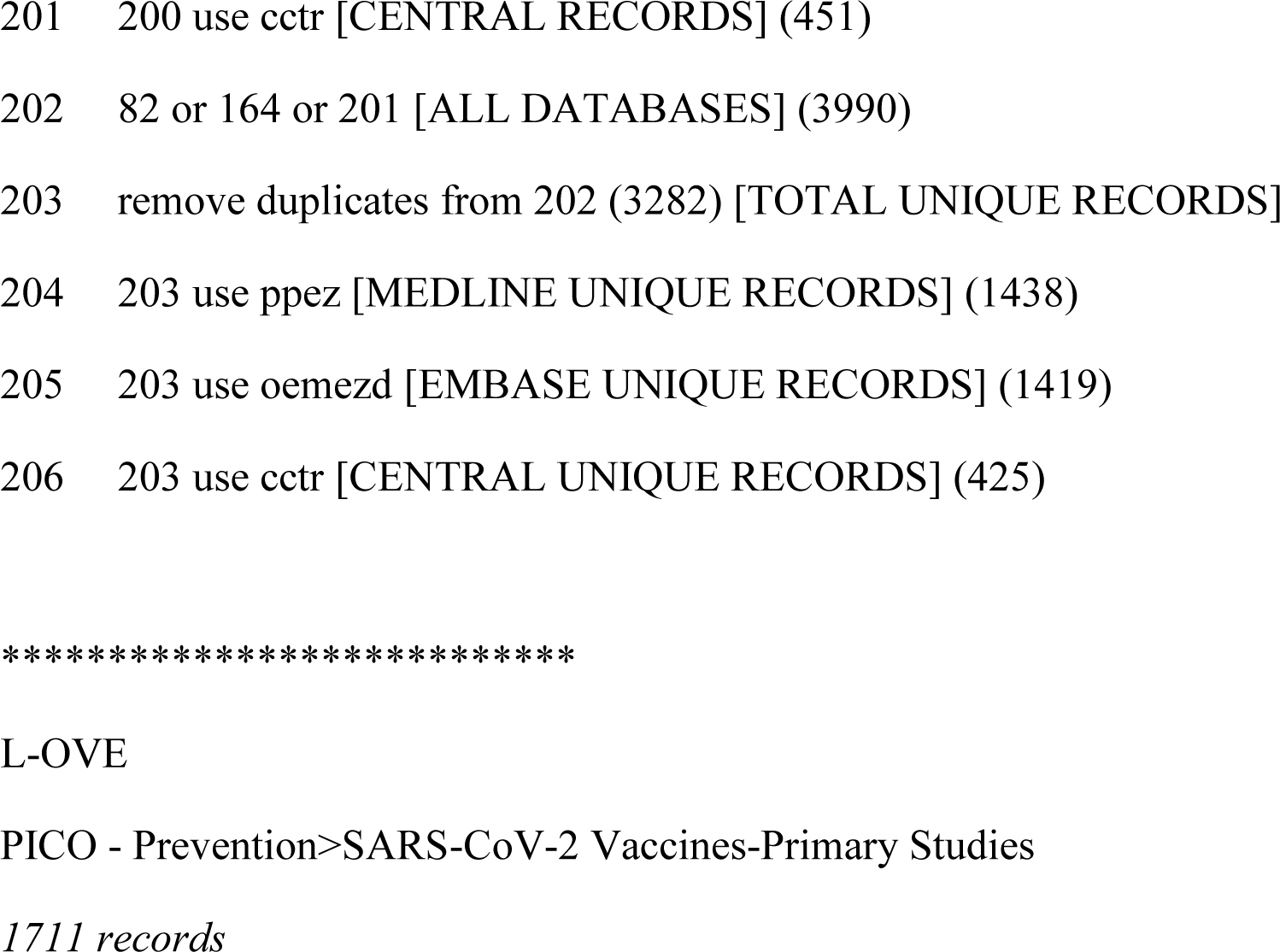

## Appendix 2: Vaccine Efficacy or Effectiveness Against Symptomatic Infection

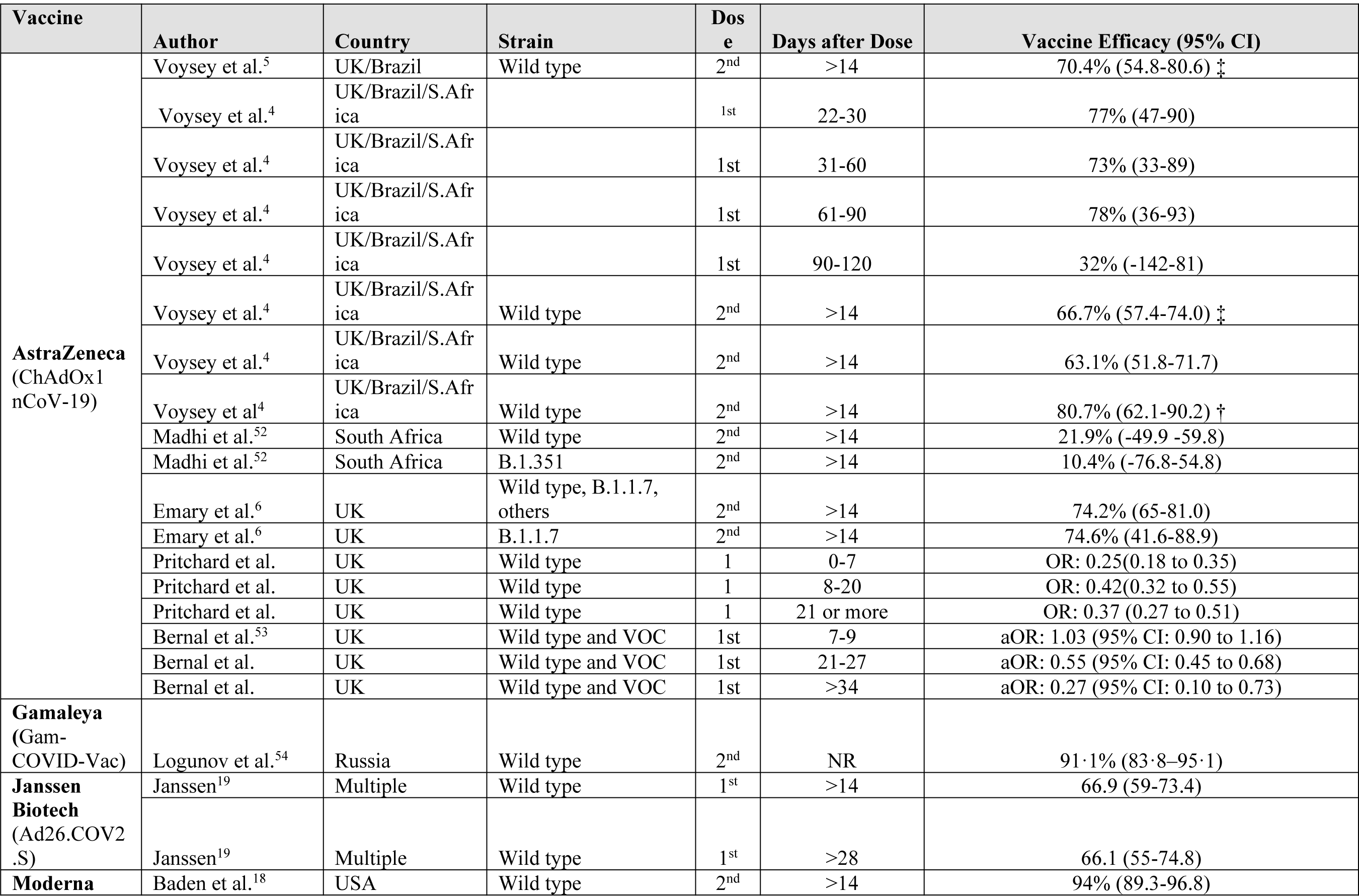
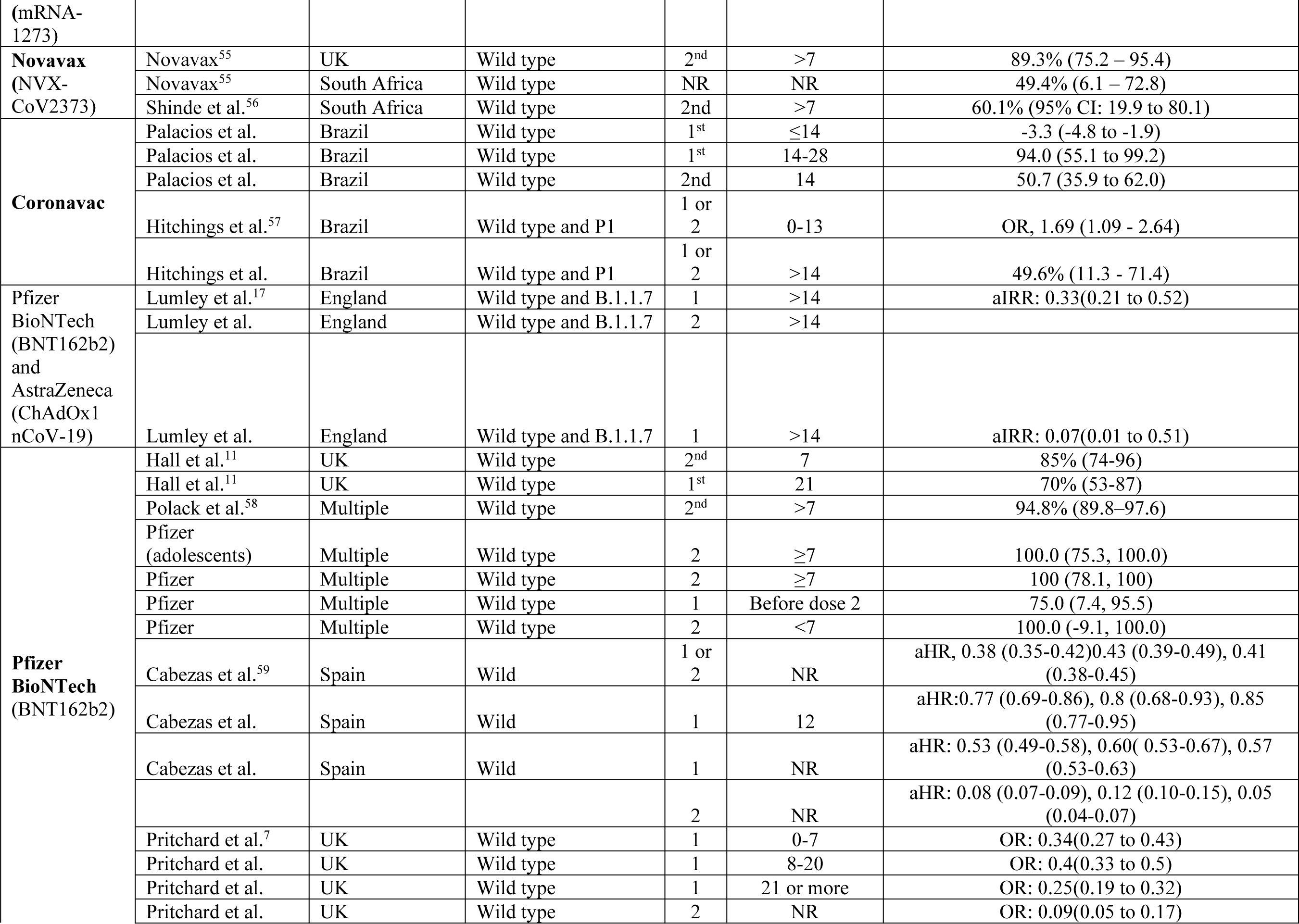
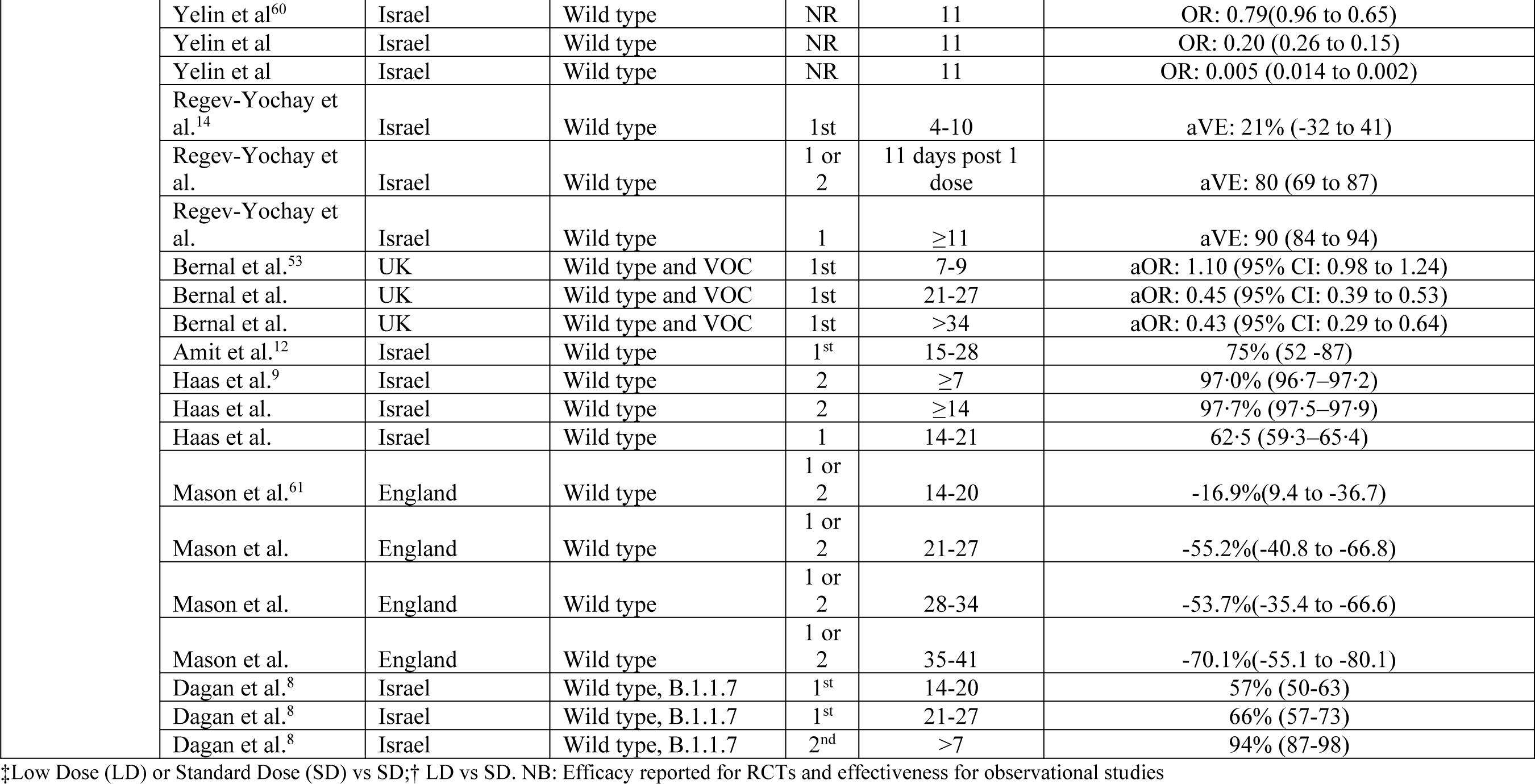

## Appendix 3: Vaccine Efficacy or Effectiveness Against Any Positive PCR (Symptomatic and Asymptomatic)

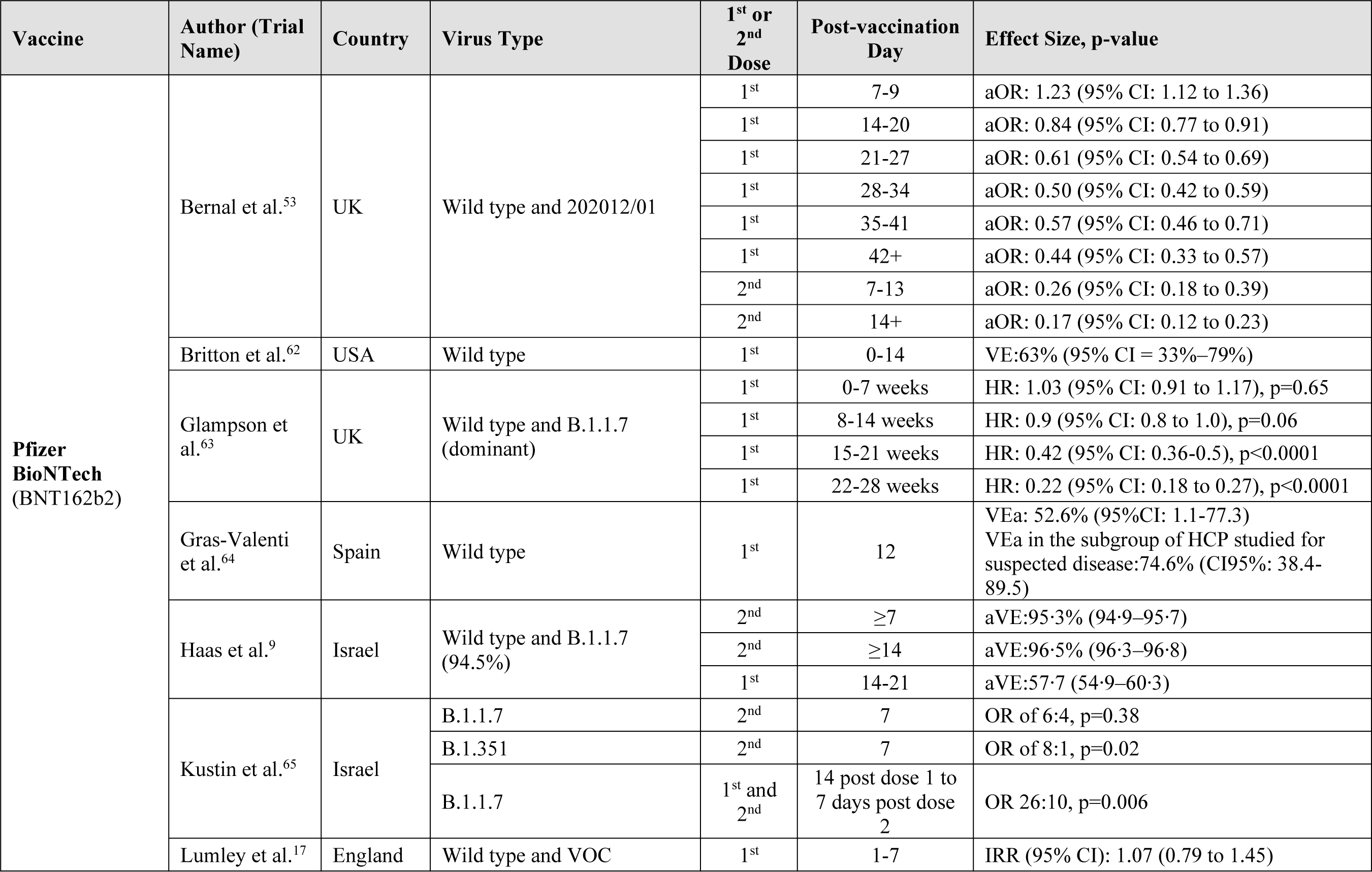
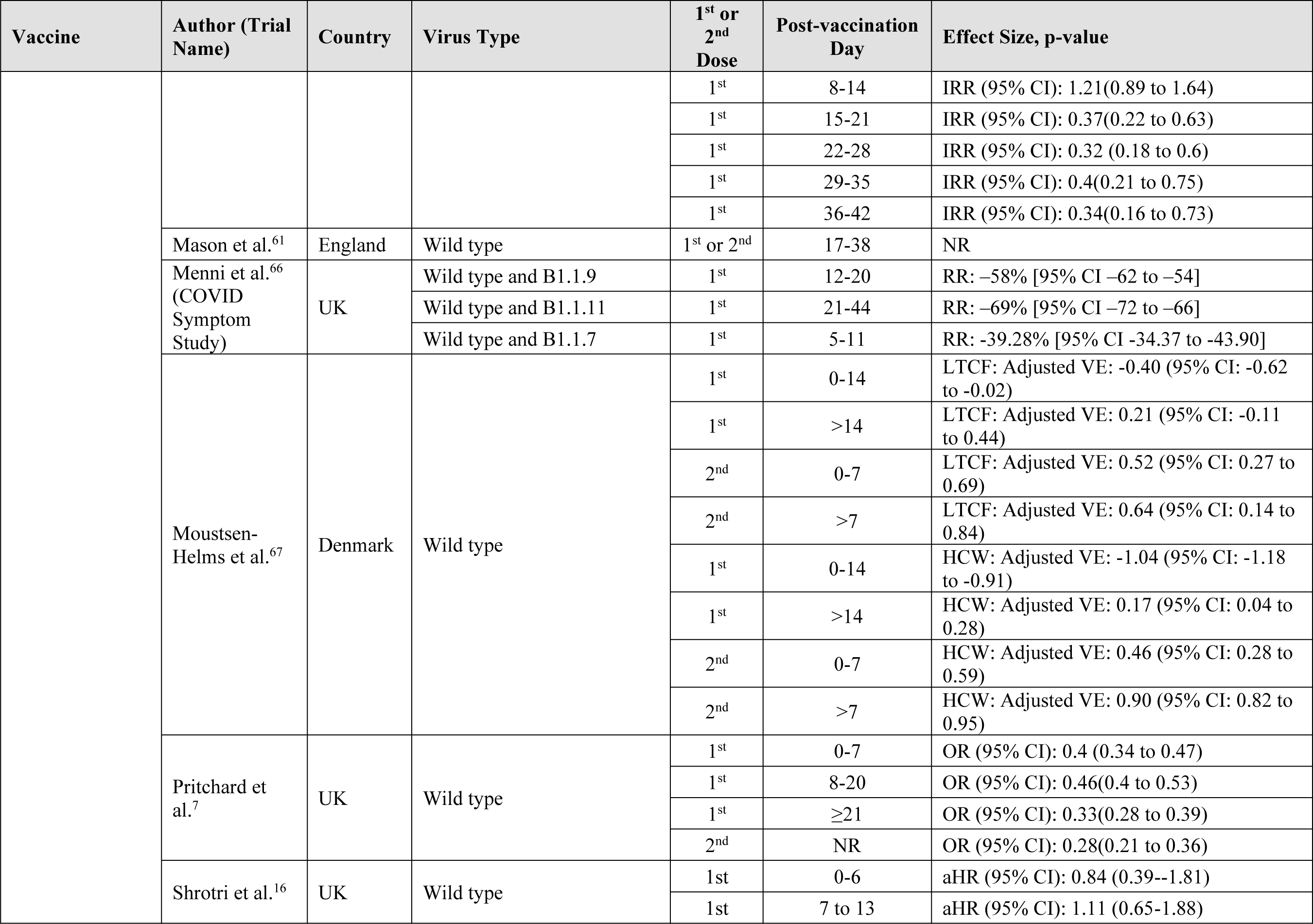
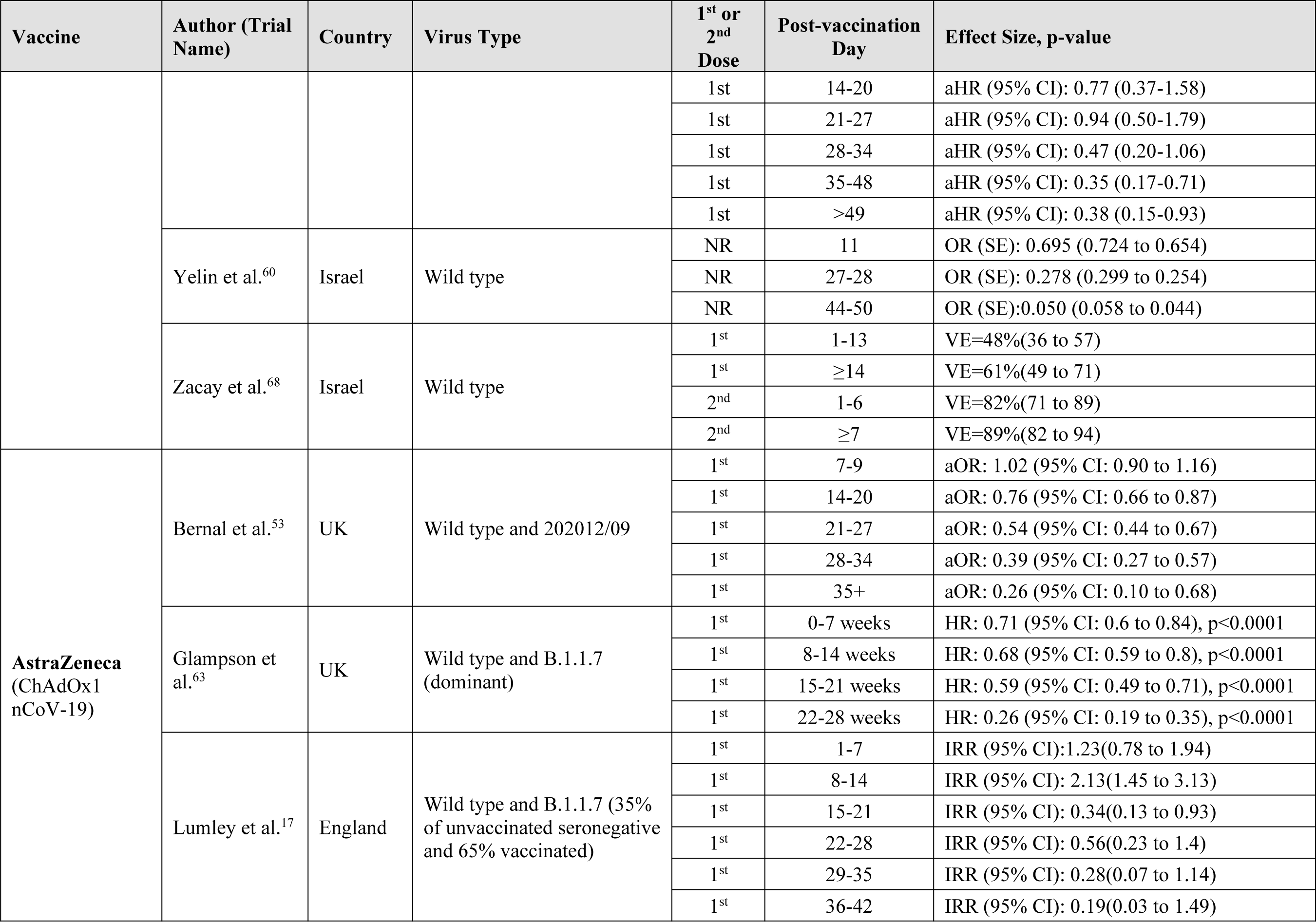
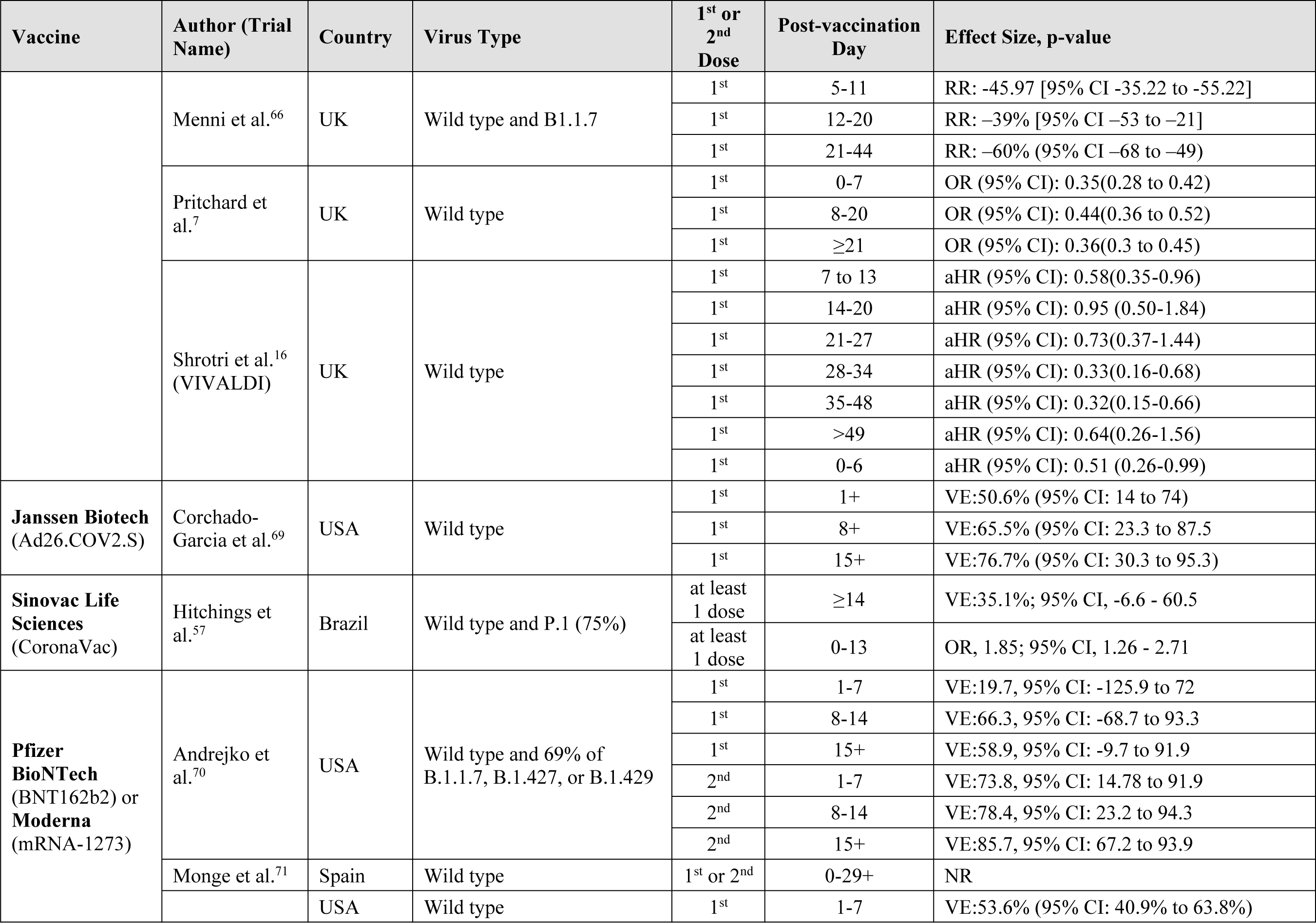
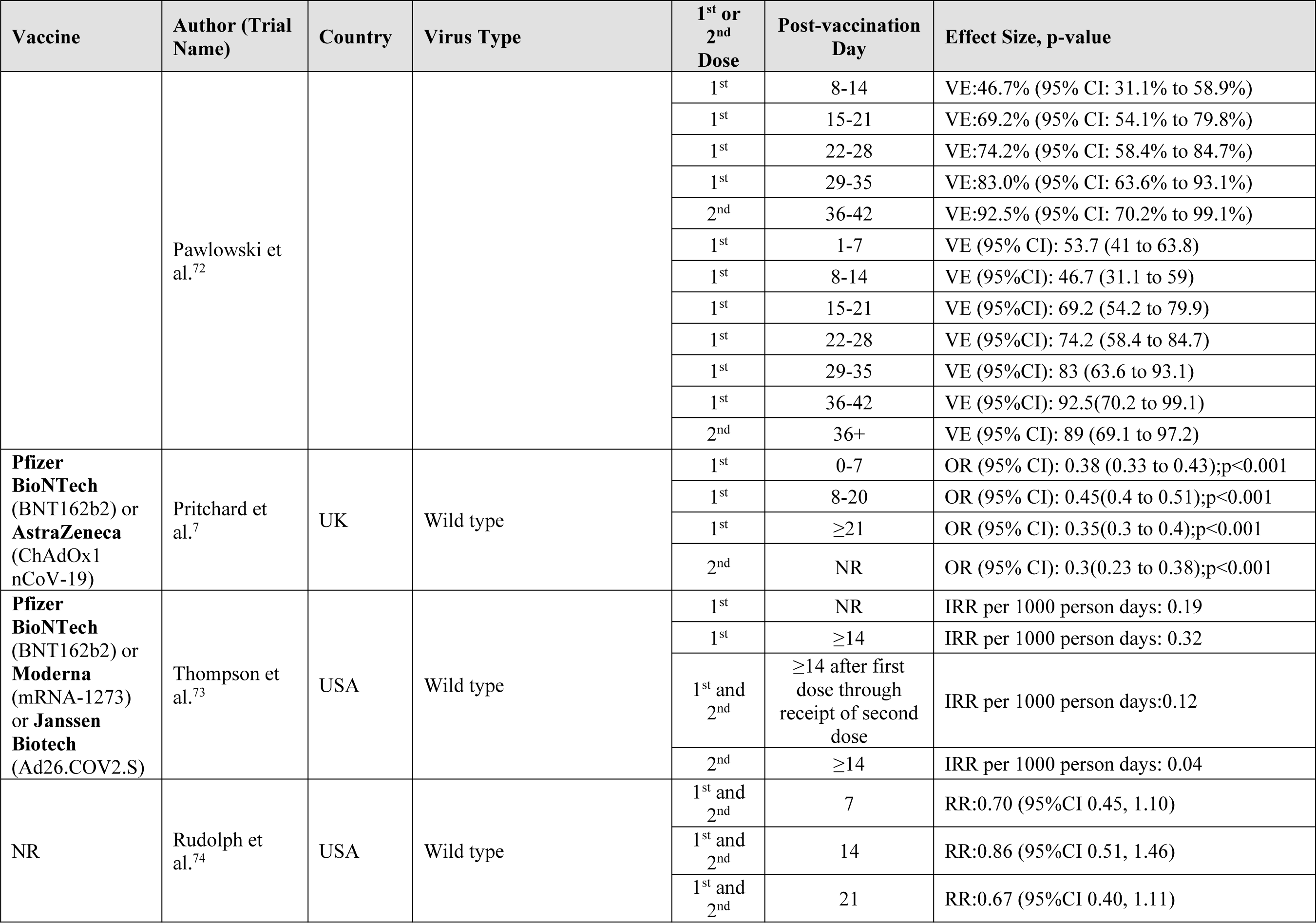
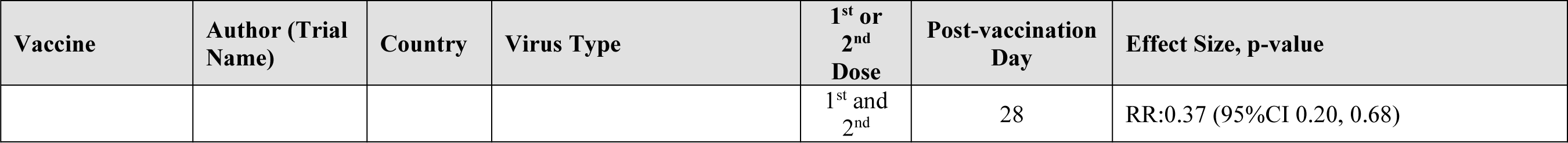

## Notes

Funding Statement: The SPOR Evidence Alliance (SPOR EA) is supported by the Canadian Institutes of Health Research (CIHR) under the Strategy for Patient-Oriented Research (SPOR) initiative. COVID-19 Evidence Network to support Decision-making (COVID-END) is supported by the Canadian Institutes of Health Research (CIHR) through the Canadian 2019 Novel Coronavirus (COVID-19) Rapid Research Funding opportunity.

### Competing Interest Statement

The authors have declared no competing interest.

### Funding Statement

The SPOR Evidence Alliance (SPOR EA) is supported by the Canadian Institutes of Health Research (CIHR) under the Strategy for Patient-Oriented Research (SPOR) initiative.
COVID-19 Evidence Network to support Decision-making (COVID-END) is supported by the Canadian Institutes of Health Research (CIHR) through the Canadian 2019 Novel Coronavirus (COVID-19) Rapid Research Funding opportunity.

